# Sequence-based variant-effect predictions reveal trait-specific regulatory-layer profiles of complex-trait heritability

**DOI:** 10.64898/2026.10.16.26365786

**Authors:** Byung Hyug Choi, Jaeseung Jeong

**Author notes:** Correspondence: Jaeseung Jeong, Ph.D. Department of Brain and Cognitive Sciences, Korea Advanced Institute of Science and Technology (KAIST), 291 Daehak-ro, Yuseong-gu, Daejeon 34141, Republic of Korea.

## Abstract

Partitioned-heritability studies have shown which tissues carry the heritability of complex traits, but whether the regulatory layer whose annotations are most informative for heritability within a tissue differs between traits has not been tested, because measured annotations for different layers come from different sets of biosamples. Sequence-to-function models predict the allelic effect of every variant on several regulatory layers with a single model, so that layers can be compared on the same variants within the same tissues. We grouped AlphaGenome Atlas predictions into 12 lineages and six regulatory layers and partitioned the heritability of 11 complex traits with stratified LD score regression. Traits localized largely to the expected lineages, and within these lineages the most informative predicted layer differed between traits (*P* = 3.6 × 10^-8^). Predicted chromatin-accessibility (DNase) effects were the most informative for height and bone mineral density, whereas predicted transcription-initiation (CAGE) effects were the most informative for Alzheimer’s disease, most brain traits and cholesterol levels. The difference persisted across sensitivity analyses, and the predicted contrast held for three related traits whose layers had been predicted in advance (*P* = 2 × 10^-7^). Predicted CAGE effects also remained informative after conditioning on fine-mapped eQTLs and measured regulatory annotations. These results identify differences across traits in the heritability information captured by predicted regulatory layers and offer testable hypotheses for investigating the regulatory basis of complex traits.

## Introduction

Most heritability of complex traits lies in non-coding variation that is thought to act through gene regulation [1,2]. Stratified LD score regression (S-LDSC) partitions SNP heritability across functional annotations using GWAS summary statistics [2,3]. Applied to tissue-specific chromatin and expression data, it and earlier enrichment methods have mapped many traits to the tissues and cell types in which their associated variants and heritability are enriched [4–6]. These maps show where a trait acts. They say less about the regulatory layer through which it acts within that tissue: whether chromatin accessibility, active histone marks, transcription initiation or steady-state expression carries the heritability.

The difficulty is partly one of measurement. Different layers have largely been profiled by different consortia in different sets of tissues and cell types. Roadmap Epigenomics measured several histone marks in the same epigenomes [5], but transcription initiation was profiled mainly by FANTOM5 in other biosamples [7,8]. A comparison between layers is therefore often also a comparison between biosample collections. Differences between traits in the most enriched mark or layer have therefore been noted descriptively [2,4,9,10], but, to our knowledge, whether the layer most informative for heritability differs between traits has not been tested formally. The question matters because GWAS signals often fail to colocalize with expression QTLs [11–13], and the cis-genetic component of assayed expression mediates only about 11% of heritability [14]. Part of the remaining signal may act through expression in contexts that have not been assayed [11], and part may act through other regulatory layers.

Sequence-to-function models offer a partial way around the measurement problem. They predict the allelic effect of every variant on many regulatory readouts with a single model and a common scoring rule, so that layers can be compared on the same variants within the same tissues [15–18]. Because the models are trained on the same consortium data, the biosamples profiled for each layer still overlap only partly; we therefore compared layers within lineages rather than within individual biosamples. Allelic-effect annotations from two earlier models, DeepSEA and Basenji, added little heritability information once baseline and measured chromatin annotations were included: only predicted H3K4me3 from Basenji remained significant, and transcription initiation, which Basenji also predicts, was left out of the analysis [19]. Applying the newest models genome-wide has been limited by computation; a recent genome-wide comparison of sequence models included AlphaGenome in variant classification but not in its heritability analysis, which was limited to models that could score about 10 million variants or had precomputed scores [20]. The AlphaGenome Atlas now provides precomputed predictions for every possible single-nucleotide variant in the human genome [21], and, to our knowledge, these predictions have not yet been used to partition heritability. In previous work we used the same predictions to examine Alzheimer’s disease (AD) loci one at a time and found that association tracked predicted expression effects, and less robustly splicing effects, but that the cellular context of the fine-mapped variants could not be resolved at the level of single loci [22]. Aggregation across the genome may provide the statistical power that single loci lack.

Here we built lineage-by-layer annotations from AlphaGenome Atlas predictions, grouping tissues and cell types into 12 lineages, and asked four questions (Fig. 1). Does heritability localize to the expected lineages? Within a lineage, does the regulatory layer whose predicted effects are most informative for heritability differ between traits? If so, does the resulting map predict the layer of traits that were not used to build it? And do the predictions carry information beyond measured annotations of matched cell types and tissues and beyond fine-mapped eQTLs?

**Figure 1.**
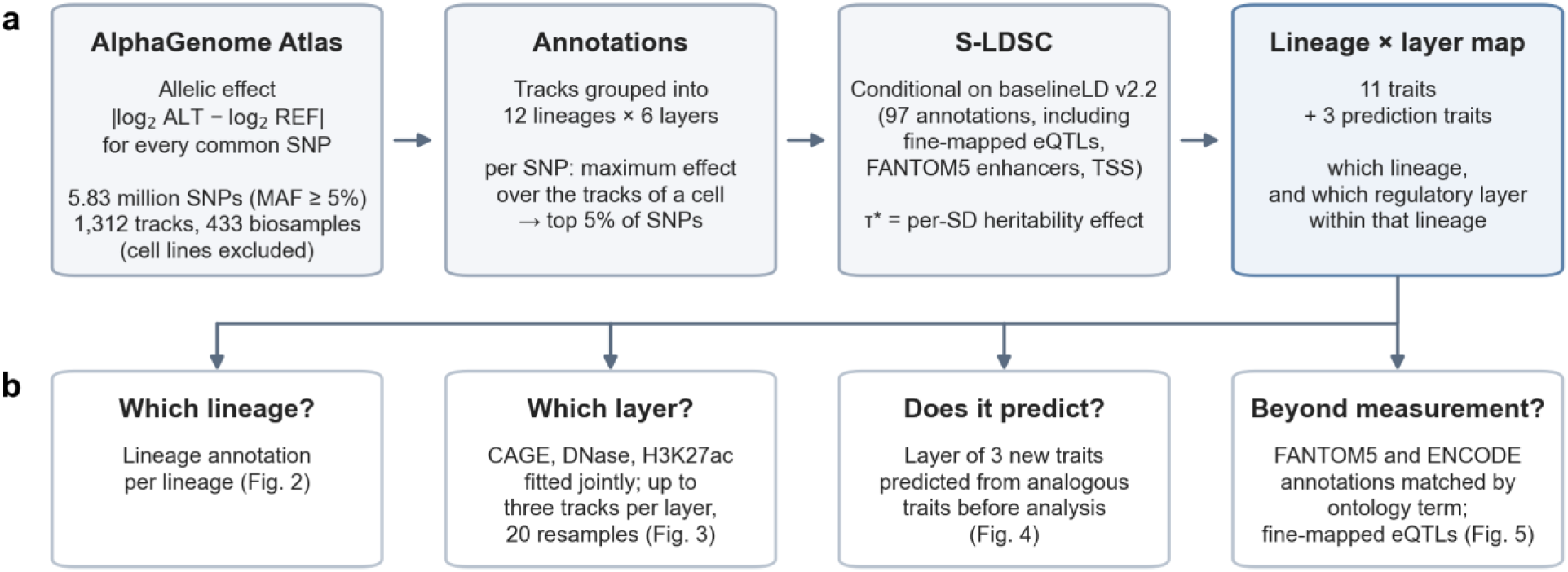
Study design. **a**, AlphaGenome Atlas predictions of the allelic effect (|log2 ALT - log2 REF|) of 5.83 million common SNPs on 1,312 tracks from 433 biosamples (cell lines excluded) were grouped into 12 lineages and six regulatory layers. For each SNP, the maximum effect over the tracks of a lineage–layer cell was ranked across SNPs, and the top 5% defined a binary annotation. Heritability was partitioned with S-LDSC conditional on the 97 annotations of baselineLD v2.2, which include fine-mapped eQTLs, FANTOM5 enhancers and promoter and TSS annotations. **b**, The four questions addressed and the figures that address them.

## Results

### Allele-specific annotations from AlphaGenome Atlas

We retrieved AlphaGenome Atlas predictions for all 5,828,323 SNPs with minor allele frequency (MAF) ≥ 5% in the 1000 Genomes European reference panel [23] for which the reference and alternative alleles could be resolved (97.9% of 5,956,110). We stored per-track allelic effects for 1,748 tracks: DNase-seq (305), CAGE (546), RNA-seq (371) and histone ChIP– seq for H3K27ac, H3K4me1 and H3K4me3 (526). After excluding cell lines (372 tracks) and 64 tracks from mixed or undefined biosamples, we assigned the remaining 1,312 tracks from 433 biosamples to 12 lineages by rules on biosample names and types that were fixed before any heritability analysis (Supplementary Table S2). The brain-tissue lineage, for example, comprised 38 adult and fetal brain regions; cultured neural cells, both in vitro differentiated (neurons and neural progenitors) and primary (astrocytes, oligodendrocyte precursors, Schwann cells and neurons), formed a separate neural-cell lineage. The myeloid lineage comprised monocytes, macrophages, dendritic cells, granulocytes, mast cells and myeloid progenitors; the Atlas contains no microglia tracks.

For each lineage and layer we took, for each SNP, the maximum absolute allelic effect across the tracks of that lineage and layer and annotated the top 5% of SNPs. We partitioned the heritability of 11 traits (Supplementary Table S1) conditional on baselineLD v2.2 and report τ*, the change in per-SNP heritability per standard deviation of the annotation, scaled by the mean per-SNP heritability [3]. Because the baseline model already contains fine-mapped eQTLs [24], FANTOM5 enhancers [8], promoters, TSS proximity and conservation, a positive τ* indicates that an annotation is informative for heritability beyond these annotations.

### Traits localize to the expected lineages

We first annotated each lineage with a single score that combined DNase, H3K27ac, H3K4me1 and CAGE predictions (Fig. 2). Of 132 trait–lineage cells, 19 were significant after correction for the 132 tests (false discovery rate < 0.05). Ten of the 11 traits had their strongest enrichment in the lineage expected from their biology. Monocyte count was enriched in the myeloid lineage (τ* = +2.82, *P* = 1 × 10^-10^), inflammatory bowel disease (IBD) and rheumatoid arthritis in the lymphoid lineage (+3.43, *P* = 5 × 10^-8^; +2.48, *P* = 3 × 10^-4^), LDL cholesterol in liver (+3.21, *P* = 4 × 10^-5^) and height in adipose, skeletal muscle, stromal (mesenchymal) and cardiovascular tissues (+1.18 to +1.77).

**Figure 2.**
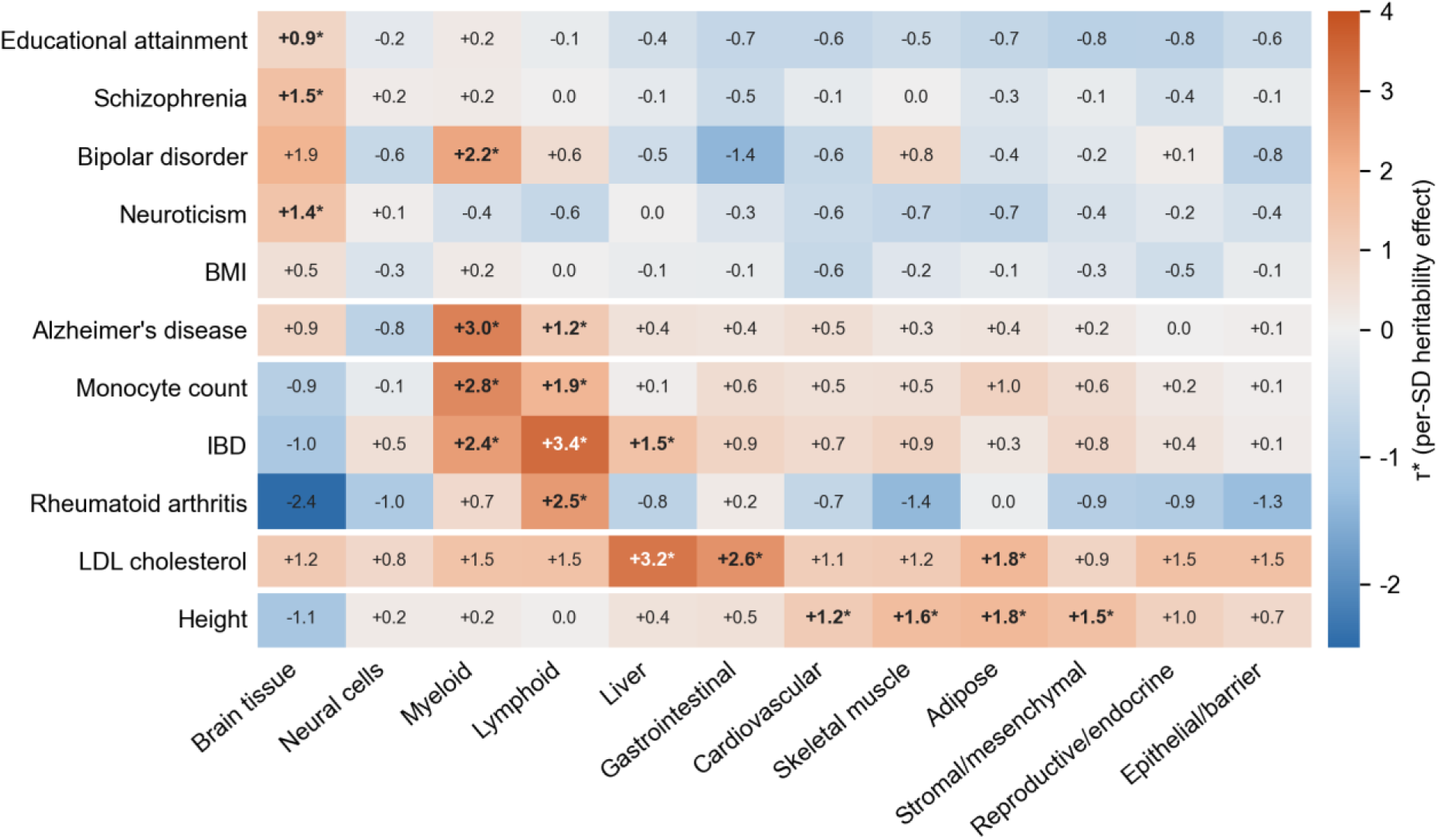
Heritability enrichment of predicted allelic effects by lineage. τ* for 11 traits (rows) and 12 lineages (columns) from models containing baselineLD v2.2 and one lineage annotation (top 5% of SNPs by the mean of the per-layer percentiles for DNase, H3K27ac, H3K4me1 and CAGE). Bold values with an asterisk are significant at false discovery rate < 0.05 across the 132 cells (one-sided test of τ* > 0). Traits are grouped as brain traits, AD, blood and immune traits, LDL cholesterol and height.

Educational attainment, schizophrenia and neuroticism were enriched in brain tissue (+0.87, *P* = 9 × 10^-5^; +1.55, *P* = 3 × 10^-9^; +1.42, *P* = 7 × 10^-5^), and body mass index (BMI) showed nominal brain-tissue enrichment (+0.53, *P* = 0.037). Bipolar disorder was the exception: its strongest enrichment was in the myeloid lineage (+2.22, *P* = 0.001), with brain tissue second (+1.91, *P* = 0.009). In contrast, the lineage of cultured neural cells was not enriched for any brain trait (τ* between −0.62 and +0.15). Brain-trait heritability was therefore captured by predictions in adult and fetal brain tissue, not by predictions in cultured neural cells. These enrichments remained when we also conditioned on an annotation of SNPs with large predicted effects in any lineage (Supplementary Fig. S1), so they reflect lineage-specific predictions rather than a general tendency of large-effect SNPs to carry heritability.

AD heritability was enriched in the myeloid lineage (+2.98, *P* = 6 × 10^-6^) and, more weakly, in the lymphoid lineage (+1.22, *P* = 0.005), in agreement with earlier evidence that AD risk acts through myeloid cells and microglia [25–28]. Enrichment in brain tissue was not detected (+0.88, *P* = 0.068). Its point estimate was similar to that of educational attainment (+0.87), but the standard error was larger, partly because AD has a low observed-scale heritability (0.026), so the result does not exclude a brain-tissue contribution.

### Within a lineage, the most informative predicted layer differs between traits

We then separated the layers within each lineage. In single-layer models (72 lineage–layer annotations per trait), 54 of 792 trait–cell combinations were significant (Supplementary Fig. S2). The strongest cell was a CAGE cell for 10 of 11 traits: brain-tissue CAGE for every brain trait, myeloid CAGE for AD, monocyte count and IBD, lymphoid CAGE for rheumatoid arthritis and liver CAGE for LDL cholesterol. For height it was stromal DNase. Predicted RNA-seq annotations were rarely informative: one of 132 RNA cells was significant. Because RNA-seq scores are gene-level, whereas CAGE and DNase scores are computed in a 501-bp window around the variant, this difference should not be read as evidence about the importance of steady-state expression.

Single-layer models cannot show which layer is most informative, because annotations of different layers in the same lineage overlap and because layers differ in the number of tracks. CAGE has the most tracks in most lineages, so its maximum-effect score could be favoured for that reason alone. We therefore fitted three distinct layers that were available in every lineage analysed, CAGE (transcription initiation), DNase (chromatin accessibility) and H3K27ac (active promoters and enhancers), jointly in the lineage relevant to each trait. In these joint models, a larger τ* for one layer means that its annotation carries more heritability information conditional on the other two layers and the baseline; it does not by itself show that the layer is the route through which risk acts. We reduced the track-number imbalance by resampling up to three units per layer (a unit was one track for DNase and H3K27ac and one CAGE library, with both strands, for CAGE; all units were used when fewer were available, as for the two liver CAGE libraries) 20 times, and averaged estimates and jackknife replicates over the 20 resamples. H3K4me1 and H3K4me3 were left out because they were collinear with H3K27ac, and RNA-seq because its gene-level annotations carried almost no signal and are not directly comparable with the window-based scores. These choices, the tests and the classification rule were fixed in a dated protocol before the joint models were run, but after the single-layer results and joint six-layer models of the same GWAS had been seen. The layer types of the 11 map traits are therefore an exploratory classification; the prospective test of the map is the prediction for three new traits described below.

The layer profiles differed between traits (multivariate Cochran’s *Q* = 74.3, 20 degrees of freedom, *P* = 3.6 × 10^-8^; *P* = 7.6 × 10^-6^ with an *F* approximation; Fig. 3). All tests between traits used a joint block jackknife that accounts for covariance between traits from shared samples and shared genetic architecture (Methods and Supplementary Table S10). For height, predicted DNase effects were the most informative: stromal DNase τ* was +1.92 ± 0.37, against +0.49 ± 0.32 for CAGE and +0.04 ± 0.31 for H3K27ac, and DNase ranked first in 19 of 20 resamples. For AD, monocyte count, educational attainment, schizophrenia, bipolar disorder, BMI and LDL cholesterol, predicted CAGE effects were the most informative. For AD, myeloid CAGE τ* was +3.22 ± 0.85, against +0.14 for DNase and +0.01 for H3K27ac, and CAGE ranked first in all 20 resamples. In the three contrasts specified in the protocol, the CAGE - DNase contrast differed between AD and height (difference +4.51 ± 1.51, *P* = 0.003) and between height and four brain traits (educational attainment, schizophrenia, bipolar disorder and neuroticism; +2.75 ± 0.58, *P* = 2 × 10^-6^). The CAGE - H3K27ac contrast was smaller in the two traits analysed in the lymphoid lineage (IBD and rheumatoid arthritis) than in the two analysed in the myeloid lineage (AD and monocyte count), but this did not reach the threshold (−1.88 ± 0.90, *P* = 0.037; α = 0.017).

**Figure 3.**
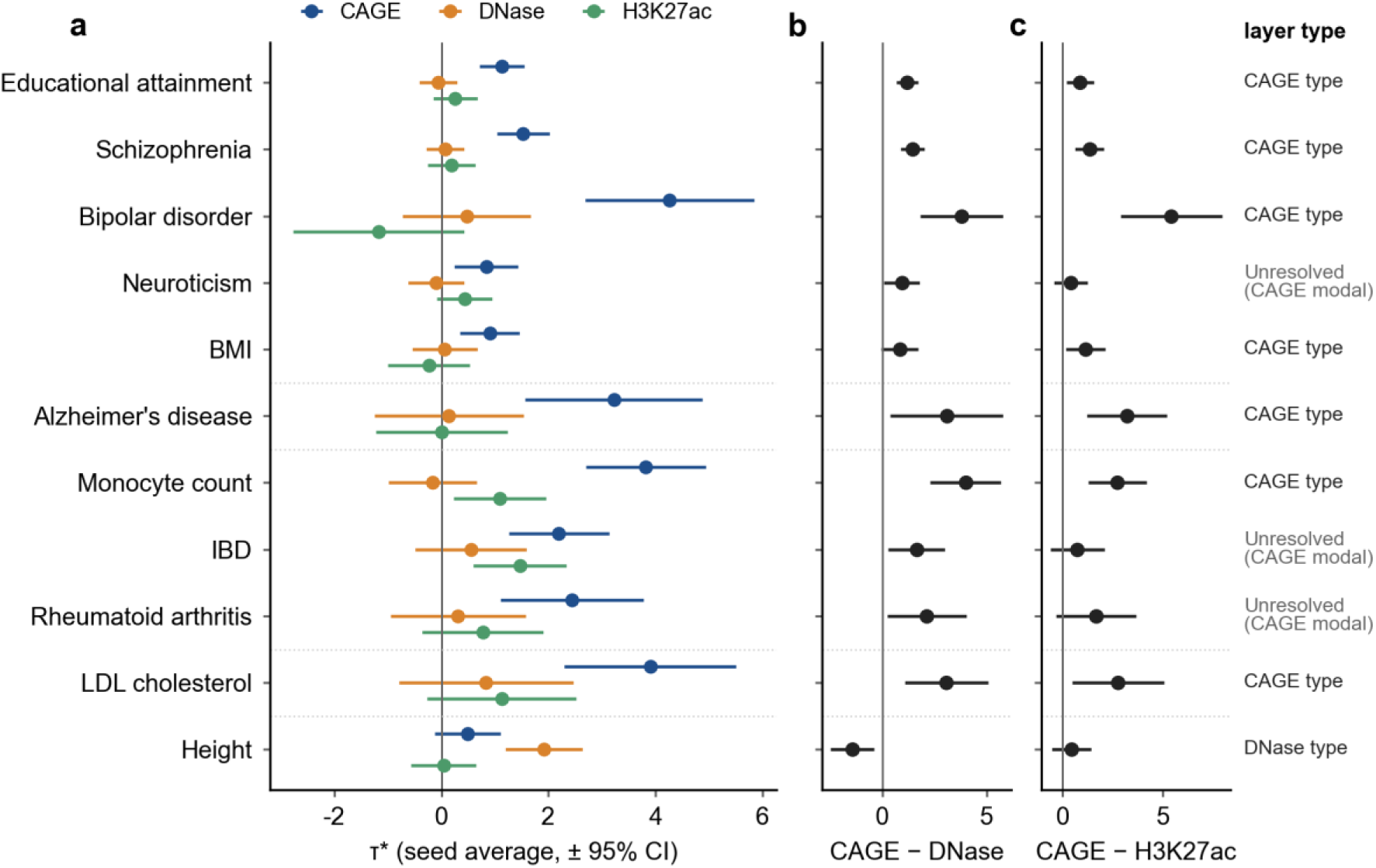
The most informative predicted layer differs between traits. **a**, τ* of predicted CAGE, DNase and H3K27ac effects fitted jointly in the lineage relevant to each trait (brain tissue for the five brain traits, myeloid for AD and monocyte count, lymphoid for IBD and rheumatoid arthritis, liver for LDL cholesterol, stromal for height). Estimates are averages over 20 resamples that limited the number of tracks (DNase, H3K27ac) or libraries (CAGE) to at most three per layer; error bars are 95% confidence intervals from the resample-averaged block jackknife. **b**, **c**, The CAGE - DNase (**b**) and CAGE - H3K27ac (**c**) contrasts for the same models, with the layer type assigned by the classification rule (a layer was called when it ranked first in the average, ranked first in at least 15 of 20 resamples and exceeded the second layer with one-sided *P* < 0.05; otherwise the trait was unresolved, and CAGE modal indicates that CAGE was the most frequent top layer). Heterogeneity across traits was tested with a multivariate Cochran’s *Q* on the two contrasts, using a joint jackknife across traits.

By the classification rule of the protocol (whose types label the annotation profile, not the mechanism), seven traits were CAGE type, height was DNase type, and three traits were unresolved (neuroticism, IBD and rheumatoid arthritis). All three unresolved traits had CAGE as the most frequent top layer, but IBD and rheumatoid arthritis ranked H3K27ac first in 4 of 20 resamples. With all tracks instead of resampled tracks, the top layer was the same for all 11 traits, so the map was not driven by track numbers. In the liver lineage, CAGE ranked first for LDL cholesterol although it had fewer units (two libraries) than DNase and H3K27ac (three tracks each). In a post hoc leave-one-trait-out analysis, heterogeneity remained after removing any single trait: *Q* was between 56 and 74 when any trait other than height was removed, and 38.5 without height (18 degrees of freedom, *P* = 0.003; Supplementary Table S5). The heterogeneity therefore has two parts: a difference in profile type between height and the other traits, and a difference in the degree of CAGE dominance among the CAGE-type traits.

In an additional analysis, we asked whether the map depends on the 5% threshold, because sequence models underestimate the effects of many causal variants and their strongest predictions are the most reliable [29]. We repeated the three-layer models with all tracks at the top 1%, 0.5% and 0.1% thresholds (Supplementary Table S7 and Supplementary Fig. S3). The criterion, fixed in a dated protocol before the analysis, was that at the 0.1% threshold layer profiles should still differ between traits (heterogeneity *P* < 0.05) and the top layer should match the main map in at least 9 of 11 traits, with height remaining DNase type. The map approached the main result as the threshold was relaxed. At the top 0.1%, where each annotation contained about 5,800 SNPs, layer profiles still differed between traits (*P* = 4 × 10^-4^), but the top layer matched the main map in only 5 of 11 traits, so the map was not reproduced by the criterion. The top layer matched in 8 of 11 traits at 0.5%, in 9 of 11 at 1% and in all 11 at 5%. At 1% this met the ranking part of the criterion, but heterogeneity between traits was not significant (*P* = 0.055). Statistical separation followed only at the broadest threshold. The contrasts of AD and the brain traits with height were significant only at 5% (at 1%, *P* = 0.057 and 0.025, against α = 0.017), and the top layer exceeded the second in 1, 0, 0 and 8 of 11 traits at 0.1%, 0.5%, 1% and 5%. The validation contrast held at every threshold (one-sided *P* = 0.018 at 0.1% and *P* ≤ 2 × 10^-4^ from 0.5% upward). At every threshold, DNase remained the top layer for height and BMD, and CAGE for AD, schizophrenia, IBD and neutrophil count. The enrichment of the top layers of AD and height increased as the threshold became stricter (for AD myeloid CAGE, from 14-fold at 5% to 129-fold at 0.1%). The largest change was in LDL cholesterol, for which DNase exceeded CAGE at 0.1% (+3.67 versus +0.54) but not at 0.5% or above. The rank order of layers is therefore largely in place from the top 1%, whereas the statistical separation of layers within and between traits required the 5% annotation.

### The layer differences persist with cell-type-matched layers, with shared annotations and at distal SNPs

Annotations of different layers within a lineage come from partly different biosamples; only 33 of the 382 biosamples (ontology terms) with any of the three layers have all three, and none in the myeloid lineage. We therefore rebuilt the three layers from matched cell types or anatomical regions, taking one unit per layer from each matched group, so that each layer had the same number of units (Supplementary Table S8). The matching table and the criterion were fixed in a dated protocol before the analysis. The top layer was unchanged for all 8 traits with a confirmed type (criterion, at least 7 of 8, including DNase for height and CAGE for AD), and CAGE ranked first for the three unresolved traits. The top layer exceeded the second in 10 of 11 traits, the layer profiles differed between traits (*P* = 3 × 10^-5^), and the contrasts of AD and the brain traits with height were significant (*P* = 6 × 10^-4^ and 3 × 10^-5^). The predicted layers of all three validation traits, including neutrophil count, were also confirmed. The layer map is therefore not explained by differences in the cell types or regions that contribute to each layer.

A second concern is that comparisons between traits used different lineages. We therefore refitted the three-layer model for each trait in every lineage in which it was enriched, so that traits could be compared with identical annotations (Supplementary Table S9). Within a lineage, track numbers, biosample composition, prediction accuracy and scorer windows are the same for all traits. In all six lineages in which both a DNase-type trait (height or BMD) and a CAGE-type trait were enriched, the CAGE - DNase contrast was larger for the lipid and blood traits than for the skeletal traits (pooled difference +2.29 ± 0.63, one-sided *P* = 1.5 × 10^-4^; *P* = 0.003 with the more conservative standard error of the protocol, which assumed perfect correlation between lineages; criterion met). The top layer itself depended on the lineage: outside the stromal and reproductive and endocrine lineages, H3K27ac ranked first for height, and outside liver it often ranked first for the lipid traits. What carries over between lineages is therefore the relative position of traits on the CAGE– DNase axis, and the layer types refer to each trait’s own lineage. AD and the brain traits were not enriched in any lineage shared with a DNase-type trait and could not be tested in this way; AD also ranked CAGE first in its second enriched lineage, lymphoid.

A third concern is that CAGE predictions may mainly reflect proximity to promoters, which current sequence models capture better than distal enhancers [30]. We therefore split each layer annotation of the main map at 1 kb from the nearest annotated transcription start site (TSS) of a protein-coding or lncRNA transcript [31] and fitted the proximal and distal parts of the three layers jointly (Supplementary Table S11). Most of each annotation was distal (81–84% for CAGE and 89–95% for DNase and H3K27ac). The criterion was fixed in a dated protocol before the analysis. At distal SNPs, CAGE remained the most informative layer for all 7 CAGE-type traits and DNase for height, the layer profiles differed between traits (*P* = 1.9 × 10^-6^), and the contrasts of AD and the brain traits with height were significant (*P* = 0.003 and 7 × 10^-5^). The predicted layers of the three validation traits were also the top distal layers. The proximal annotations, which covered only 0.2–1.0% of SNPs and had large standard errors, did not separate the traits (*P* = 0.54). The CAGE type therefore does not reflect promoter proximity alone.

### The layer map predicts the layer of new traits

To test whether the map predicts, we chose three traits that were not used to build it and fixed predictions in a dated protocol before analysing them. Each new trait was paired with an analogous trait already in the map, and its predicted layer was set to the layer type of that analogue: heel bone mineral density (BMD) [32] from height (stromal lineage, DNase type), total cholesterol from UK Biobank [33] from LDL cholesterol (liver, CAGE type), and neutrophil count [34] from monocyte count (myeloid, CAGE type). The BMD and total cholesterol GWAS share no samples with their analogues, whose GWAS preceded UK Biobank [35,36]. Neutrophil count and monocyte count come from the same study and are therefore a weaker test.

The predicted cross-trait contrast held (Fig. 4). The inverse-variance-weighted CAGE - DNase contrast was +2.21 for the two traits predicted to be CAGE type and −1.21 for the trait predicted to be DNase type (difference +3.42 ± 0.67, one-sided *P* = 1.6 × 10^-7^). BMD was DNase type, as predicted: stromal DNase τ* was +1.92, DNase ranked first in all 20 resamples, and it exceeded CAGE (*P* = 0.004) and H3K27ac (*P* = 0.003). Total cholesterol was CAGE type, as predicted: liver CAGE τ* was +3.70, ranked first in all 20 resamples, and exceeded DNase (*P* = 7 × 10^-5^) and H3K27ac (*P* = 0.004). Neutrophil count ranked CAGE first in 18 of 20 resamples and CAGE exceeded DNase (*P* = 0.008), but it was not distinguished from H3K27ac (*P* = 0.085). By the pre-specified criterion, which required the predicted layer to exceed both others, this prediction was not confirmed. The predicted lineages were enriched for all three traits (Supplementary Table S1): myeloid for neutrophil count (τ* = +2.04, *P* = 2 × 10^-11^, first of 12 lineages), liver for total cholesterol (+1.97, *P* = 0.002, first), and stromal for BMD (+1.85, *P* = 2 × 10^-10^, third after adipose and skeletal muscle, the same mesenchymal pattern as height).

**Figure 4.**
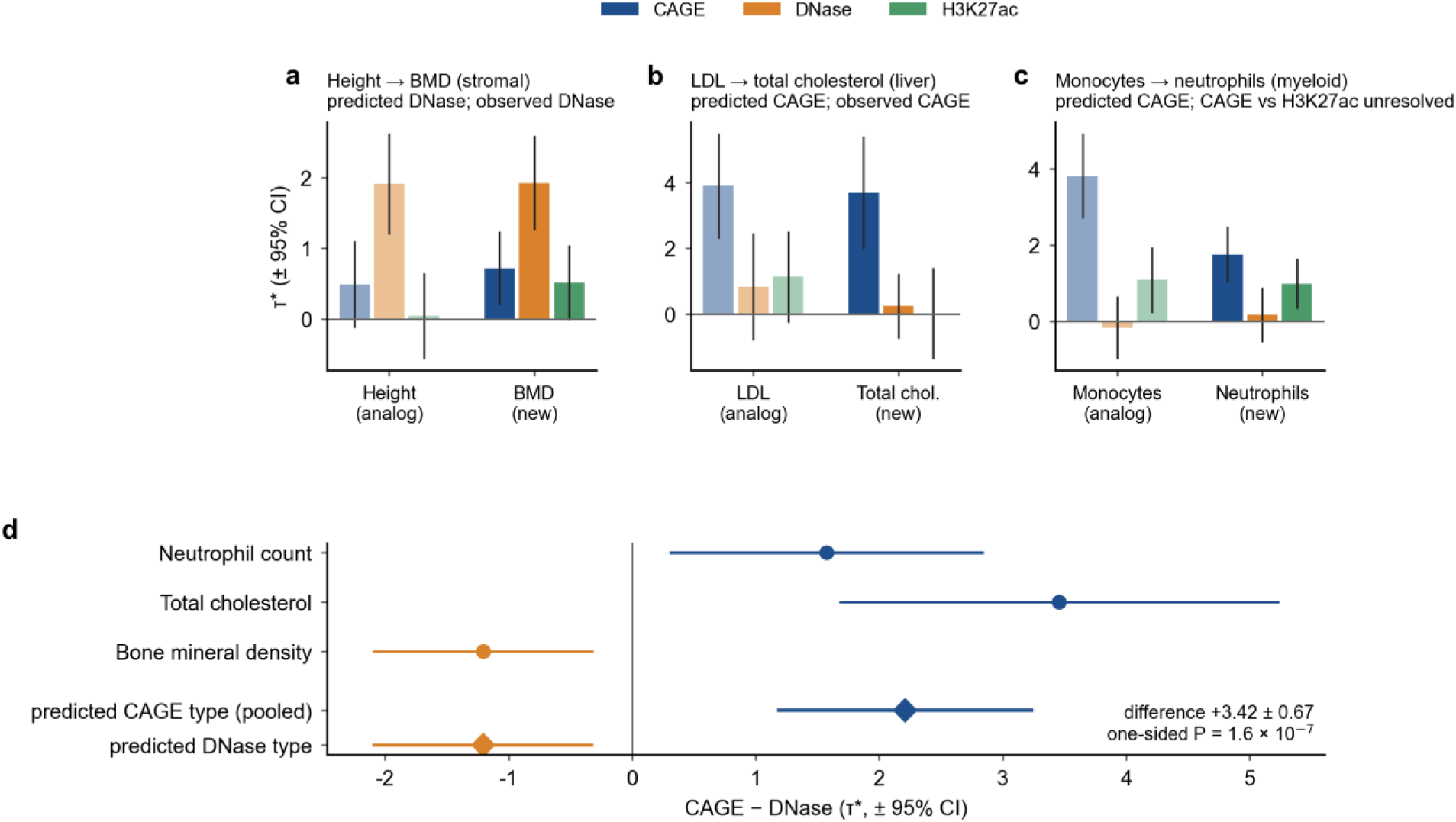
Pre-specified prediction of layer type for three new traits. **a–c**, Layer profiles (τ*, three layers fitted jointly, averaged over 20 resamples; 95% confidence intervals) of each analogous trait from the map (light bars) and the new trait (dark bars). The predicted lineage and layer were fixed in a dated protocol before the new traits were analysed. **d**, The CAGE - DNase contrast for the three new traits and the inverse-variance-weighted means of the traits predicted to be CAGE type (neutrophil count, total cholesterol) and DNase type (BMD). The primary test was the one-sided difference between these means, with a joint jackknife across traits.

### Predicted annotations remain informative after conditioning on eQTLs and measured annotations

We next asked whether the predicted annotations carry heritability information beyond measured data. The question is whether a predicted annotation keeps a positive τ* when measured annotations are in the same model, not whether its τ* is larger than theirs, because τ* depends on the size and distribution of an annotation. The baseline model already contains a fine-mapped eQTL annotation pooled over 44 GTEx tissues [24,37], so every τ* in this study is conditional on it. We also built measured annotations for the same ontology terms from the data sources on which AlphaGenome was trained: FANTOM5 CAGE peaks and transcribed enhancers for CAGE [7,8], and ENCODE DNase-seq and H3K27ac peaks [38], with windows matched to the scorer windows of the predictions (Methods). These annotations mark positions where a regulatory element is active in the relevant tissue. They covered 0.5–1.2% (CAGE), 5–23% (DNase) and 12–17% (H3K27ac) of common SNPs, against 5% for the predicted annotations.

In the primary test, specified before the comparison was run, the predicted CAGE annotation of each trait’s lineage was fitted together with the measured CAGE, DNase and H3K27ac annotations of the same lineage. It remained significant in 8 of 11 traits (false discovery rate < 0.05; Supplementary Table S4); the exceptions were neuroticism, BMI and height. For AD, myeloid CAGE τ* was +2.17 ± 0.70 (*P* = 0.001) after conditioning. Only about half of the SNPs in the predicted CAGE annotations fell within any measured active window. The remaining information is therefore not explained by the position of a measured active element. Whether it reflects the predicted allelic effect itself or predicted activity outside measured peaks was not tested.

After the layer map was known, we repeated the comparison at each trait’s own layer, which was DNase for height and CAGE for the other ten traits (post hoc; Fig. 5 and Supplementary Table S3). This reused existing models except for one new conditional model for height. Conditional on the three measured layers, the predicted own-layer τ* remained positive in 10 of 11 traits (one-sided *P* < 0.05, uncorrected) and retained a median of 71% of its unconditioned value (Fig. 5a). For height, stromal DNase τ* was +1.27 (*P* = 9 × 10^-4^) after conditioning, whereas stromal CAGE, the annotation of the primary test, was not informative (+0.07, *q* = 0.42).

**Figure 5.**
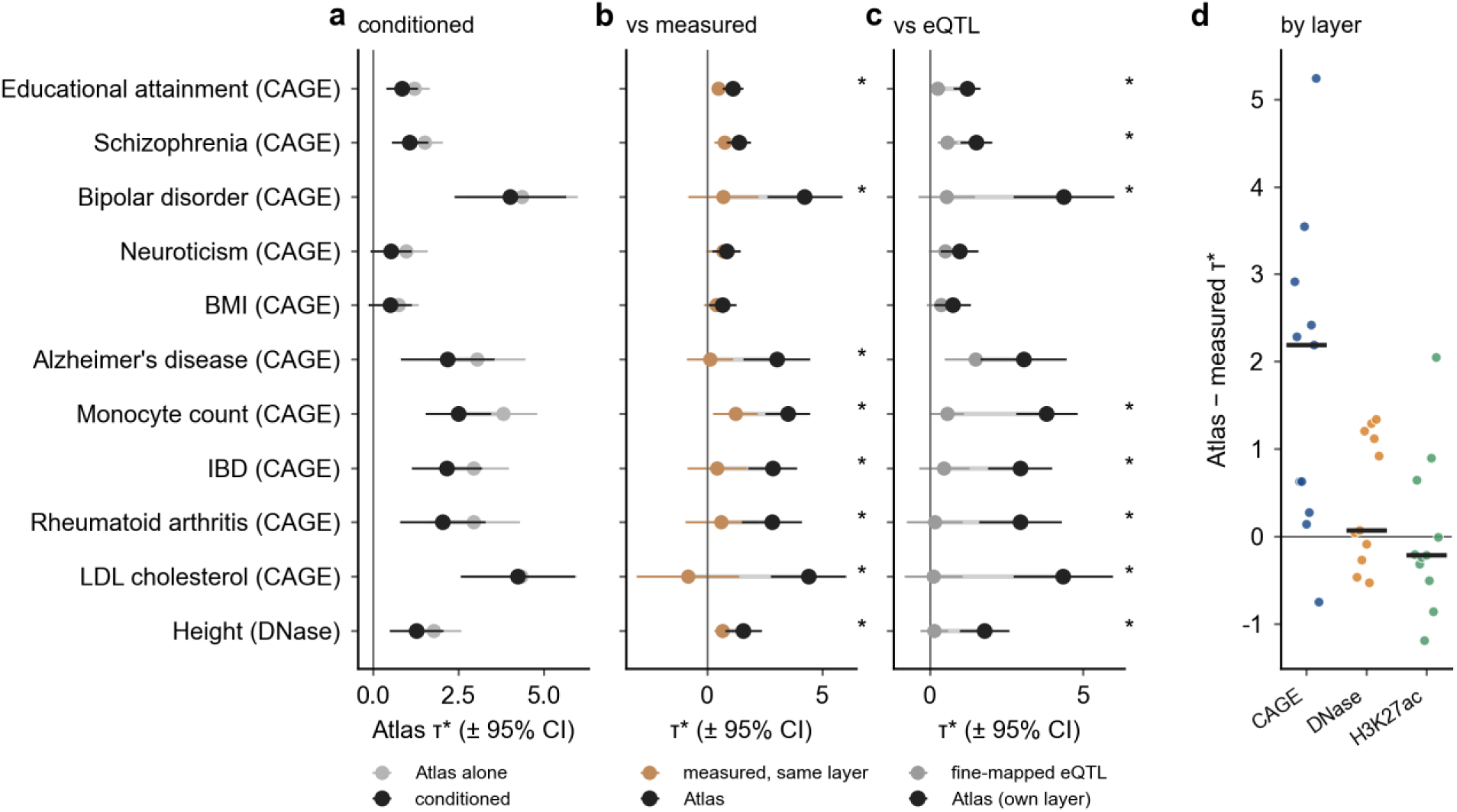
Predicted effects remain informative after conditioning on measured annotations and fine-mapped eQTLs. Each trait is shown at its own layer from the layer map (in parentheses), which was chosen after the layer map was known (post hoc); the primary test, with predicted CAGE for every trait, is in Supplementary Table S4. **a**, τ* of the predicted annotation alone and after conditioning on measured CAGE, DNase and H3K27ac annotations of the same lineage. **b**, τ* of the predicted and the measured annotation of the same layer, fitted together; measured annotations were built from FANTOM5 (CAGE) or ENCODE (DNase, H3K27ac) data for the same ontology terms as the Atlas tracks. **c**, τ* of the predicted own-layer annotation and of the fine-mapped GTEx eQTL annotation (MaxCPP) in the same model. In **b** and **c**, asterisks mark differences with two-sided *P* < 0.05, uncorrected for multiple testing. **d**, Predicted minus measured τ* for each layer across the 11 traits (points) with the median (bar). Measured annotations covered 0.5–1.2% (CAGE), 5–23% (DNase) and 12– 17% (H3K27ac) of common SNPs, against 5% for the predictions, so these differences partly reflect annotation size. Error bars are 95% confidence intervals.

Two further comparisons are descriptive. When the predicted and measured annotations of the same layer were fitted together, the predicted annotation had the larger τ* in all 11 traits and the measured annotation was never significantly larger (Fig. 5b), and adding the predicted annotation reduced the measured τ* by a median of 29% for CAGE (80% for AD), 5% for DNase and 2% for H3K27ac. The predicted-minus-measured τ* was largest for CAGE (median +2.20) and smallest for H3K27ac (−0.21; Fig. 5d). Because the measured CAGE annotations were the smallest and the measured H3K27ac annotations the largest, and τ* scales with the standard deviation of an annotation for a given per-SNP effect, these differences partly reflect annotation size and are not a measure of relative performance. Similarly, the predicted own-layer τ* was larger than the eQTL τ* in all 11 traits (Fig. 5c), but the two annotations differ in definition and distribution. Adding the predicted annotation reduced the eQTL τ* by a median of 18%.

### Predicted myeloid CAGE effects are the most informative annotation for Alzheimer’s disease

Among the brain-related traits, AD stood out. Its heritability localized to the myeloid lineage rather than to brain tissue, and within the myeloid lineage predicted CAGE effects were the most informative. This signal was robust across the analyses in Table 1. In an initial analysis with a narrower monocyte and macrophage annotation, the enrichment remained significant at the top 1% and top 10% thresholds and with a continuous annotation. It remained after conditioning on the overall AlphaGenome variant-impact score and after including the *APOE* region. In a post hoc analysis that removed 162 Mb within 1 Mb of genome-wide significant loci, τ* retained 68% of its value (+1.88, *P* = 0.0015), and no single jackknife block reduced τ* below +2.46. The myeloid signal is therefore not driven by a few large loci. In the lineage-specific model, conditional on large predicted effects in any lineage, the myeloid τ* increased to +3.82. Within the myeloid lineage, CAGE ranked first in all 20 resamples, and the CAGE annotation retained 71% of its τ* after conditioning on measured myeloid CAGE, DNase and H3K27ac annotations.

**Table 1.** Robustness of the myeloid enrichment of Alzheimer’s disease heritability. τ* with standard errors and one-sided *P* values for τ* > 0, all conditional on baselineLD v2.2 and with the *APOE* region excluded unless stated. Rows marked † were specified after the main results were seen.

| Analysis | Annotation | $\tau^*$ (s.e.) | $P$ |
| --- | --- | --- | --- |
| Lineage map (Fig. 2) | Myeloid lineage, top 5% | +2.98 (0.68) | $6.4 \times 10^{-6}$ |
| Lineage-specific model | Myeloid lineage, conditional on all-lineage annotation | +3.82 (0.92) | $1.6 \times 10^{-5}$ |
| Single layer | Myeloid CAGE | +3.05 (0.72) | $1.0 \times 10^{-5}$ |
| Layer map (Fig. 3) | Myeloid CAGE, joint with DNase and H3K27ac, 20 resamples | +3.22 (0.85) | first in 20/20 |
| Measured conditioning (Fig. 5) | Myeloid CAGE, conditional on measured CAGE, DNase and H3K27ac | +2.17 (0.70) | $9.5 \times 10^{-4}$ |
| Initial analysis | Monocyte and macrophage tracks, top 5% | +2.75 (0.65) | $1.3 \times 10^{-5}$ |
| Threshold | top 1% | +3.13 (0.91) | $2.8 \times 10^{-4}$ |
| Threshold | top 10% | +2.33 (0.54) | $8.3 \times 10^{-6}$ |
| Threshold | continuous score | +1.39 (0.41) | $3.5 \times 10^{-4}$ |
| Conditional on overall variant impact | top 5%, conditional on AVI top 5% | +2.67 (0.67) | $3.6 \times 10^{-5}$ |
| APOE region included | top 5% | +2.83 (0.67) | $1.2 \times 10^{-5}$ |
| Loci removed† | top 5%, $\pm 1$ Mb of genome-wide significant loci removed (162 Mb) | +1.88 (0.63) | $1.5 \times 10^{-3}$ |
| Leave one block out† | top 5%, minimum over 200 blocks | +2.46 | all blocks > 0 |
| Lineage map (Fig. 2) | Brain-tissue lineage, top 5% | +0.88 (0.59) | 0.068 |

## Discussion

By aggregating sequence-model predictions across the genome, we obtained a map of complex-trait heritability with two axes. The first axis, the lineage, largely reproduced known biology: brain traits mapped to brain tissue, immune and blood traits to lymphoid and myeloid cells, LDL cholesterol to liver and height to mesenchymal tissues. The second axis, the most informative predicted layer within the lineage, differed between traits. At the main threshold, this difference was significant after differences in track numbers were reduced and covariance between traits was accounted for, and for three traits analysed only after their layer had been predicted, the predicted layer was confirmed in two and the predicted contrast between them held. The difference persisted when the three layers were built from matched cell types, when only SNPs more than 1 kb from annotated TSSs were considered and, for skeletal versus lipid and blood traits, when traits were compared with identical annotations.

Earlier work reported that traits differ in the chromatin marks or layers that are most enriched for their associations [2,4,9,10]. Those comparisons largely combined marks measured in different sets of biosamples and were descriptive. Predicted effects on several layers for the same variants and lineages allowed us to fit the layers jointly, reduce differences in their track numbers and test the trait-by-layer interaction formally. The lineage axis, the myeloid localization of AD and the mesenchymal localization of height are not new; tissue-specific scores from earlier sequence models are also enriched for heritability in relevant traits [20]. What is new, to our knowledge, is a formal test of the layer axis and evidence that a trait’s layer profile carries over to related traits. Our results also refine those of Dey et al. [19], who found that deep-learning allelic-effect annotations added little beyond baseline annotations, with predicted H3K4me3, a promoter-associated mark, as the exception. We found substantial conditional signal for predicted transcription initiation, a layer that their analysis did not include. Both results suggest that the information that sequence models add beyond annotation databases may be concentrated near sites of transcription initiation.

The two profile types are hypotheses about regulatory routes, not demonstrations of them. A larger conditional τ* can reflect biology, but it can also reflect differences between assays in prediction accuracy, in the quality of the training data, in signal near promoters and in how scorers aggregate signal. With that caveat, skeletal growth and bone formation depend on mesenchymal differentiation programmes, and height heritability is enriched in accessible chromatin of growth-plate chondrocytes [39,40]. In measured data, active histone marks of chondrogenic cells are at least as strongly enriched [2], so our result concerns predicted allelic effects on accessibility rather than the location of accessible regions. This would fit a trait in which risk acts partly through enhancer accessibility. For most brain, blood and lipid traits, the most informative annotation was predicted transcription initiation, which occurs at promoters and enhancers [8,41,42]. This held at SNPs more than 1 kb from annotated TSSs, which is consistent with, but does not demonstrate, a contribution from enhancer transcription. Genetic effects on promoter usage are context specific and contribute to complex traits [43,44], and predicted CAGE annotations captured heritability not accounted for by fine-mapped eQTLs. Transcription initiation is therefore a candidate layer for part of the regulation that eQTL studies miss [11–13]. We regard this as an interpretation and have not tested it directly.

The difference between predicted and measured annotations also varied by layer: it was largest for CAGE and absent for H3K27ac. Part of this pattern reflects annotation size, because τ* scales with the standard deviation of an annotation and the measured CAGE annotations were the smallest and the measured H3K27ac annotations the largest. A second explanation is the resolution of each readout. Histone acetylation marks regions of several kilobases, so knowing where a marked region lies may capture most of its information. Transcription initiation depends on sequence features at base-pair resolution, such as core promoter elements and the initiation sites of enhancer RNAs, so the effect of a single base change can differ even within an active element. Differences in prediction accuracy between layers and in the scoring windows (501 bp for CAGE and DNase, 2,001 bp for histone marks) could also contribute, and we did not distinguish these explanations. If the resolution explanation holds, sequence models may be most useful where the regulatory readout is resolved at the base-pair level.

For AD, the most informative annotation was predicted transcription initiation in myeloid cells. The myeloid localization agrees with earlier epigenomic and expression studies [25–28,45,46]. The layer assignment is, to our knowledge, new. Brain-tissue enrichment was not detected, but this is not evidence against it. AD has a low observed-scale heritability, the point estimate was similar to that of educational attainment, and a recent analysis of brain single-cell data found no significant AD enrichment in any brain cell type, including microglia, under conservative conditioning [47]. Our myeloid signal may differ from that result because the Atlas myeloid lineage consists of peripheral myeloid cells and because predicted allelic effects differ from measured peaks. Because the Atlas contains no microglia tracks, the myeloid lineage stands in for microglia only through shared myeloid programmes, which are enriched for AD heritability to a similar degree in monocytes, macrophages and microglia [28].

The map also connects with our locus-level study [22]. Within single AD loci, the same predictions did not resolve a shared cellular context for the fine-mapped variants. Genome-wide, the lineage and layer were resolved. The cellular and layer information in sequence-model predictions is therefore more informative in aggregate than for interpreting individual loci, where fine-mapping uncertainty and allelic heterogeneity may dominate.

Our study has limitations. First, enrichment of heritability locates where predicted effects co-occur with heritability; it does not identify causal cell types or mechanisms. Second, we analysed common SNPs in European-ancestry GWAS, several of them older and smaller (for example, bipolar disorder, *N* = 16,731). Third, our main analyses used binary annotations of the top 5% of SNPs. The rank order of layers was largely retained down to the top 1%, but heterogeneity between traits was not significant at 1%, the pairwise contrasts were significant only at 5%, and at the top 0.1% the top layer matched the main map in only 5 of 11 traits. The top layers retained at every threshold were DNase for height and BMD and CAGE for AD, schizophrenia, IBD and neutrophil count. Fourth, current sequence models capture promoter determinants of expression better than distal enhancers [30]. The layer map held at SNPs more than 1 kb from annotated TSSs, but distal CAGE signal may come from unannotated promoters as well as from enhancer RNAs, which our analysis cannot distinguish, and the proximal annotations were too small to compare layers. Fifth, layers within a lineage come from partly different biosamples, and comparisons between traits used different lineages. Cell-type-matched annotations and comparisons within shared lineages argue against both explanations, but matching was at the level of cell type or region rather than donor, laboratory or developmental stage; AD and the brain traits could not be compared with DNase-type traits in a shared lineage; and the top layer itself depended on the lineage. Sixth, the lineage of each of the 11 map traits was chosen after the lineage map had been seen, and the layer-map protocol was written after the single-layer results; the layer types of the map traits are therefore exploratory. For the three validation traits, the lineage and layer were predicted in advance; the lineage was confirmed in all three and the layer in two. Seventh, only two layer types emerged, and no trait was classified as H3K27ac type. The accessibility type rests on a single map trait, height, and on the related trait BMD. Without height the heterogeneity was smaller, and it reflected differences in the degree of CAGE dominance (post hoc). Whether other traits are of accessibility type remains to be tested. Eighth, several traits share UK Biobank samples or genetic architecture. A joint block jackknife over genomic blocks aligned across traits, which accounts for the resulting covariance, left all conclusions unchanged, and error correlations between traits were small (median +0.03; largest +0.39, for monocyte and neutrophil counts). The BMD and total cholesterol GWAS shared no samples with their analogues, whereas neutrophil and monocyte counts came from the same study, and all three validation traits are biologically related to their analogues. The validation therefore shows that a layer type carries over to a related trait, not that it holds for unrelated traits. Finally, Atlas coverage is uneven across lineages; for example, the myeloid lineage has four DNase tracks.

In summary, sequence-based predictions of allelic effects, aggregated genome-wide, show that complex traits differ not only in the tissue but also in the regulatory layer whose predicted effects are most informative for their heritability. They can also predict the layer of related traits, and in most traits they carry information beyond measured annotations of matched cell types and tissues. Such a map can guide the choice of follow-up assays for a trait, for example accessibility QTLs for an accessibility-type trait and CAGE, PRO-cap or reporter assays for a transcription-initiation-type trait, and it can supply layer-specific priors for fine-mapping.

## Methods

### GWAS summary statistics

We analysed 11 traits for map construction and three traits for validation (Supplementary Table S1). AD summary statistics were from stage I of the European Alzheimer & Dementia Biobank meta-analysis (*N* = 487,511, including proxy cases) [48]; the *APOE* region (chr19:43.9– 45.9 Mb, GRCh38) was removed. Monocyte count [34] and the validation traits, heel BMD [32], neutrophil count [34] and serum total cholesterol in UK Biobank [33], were obtained from the GWAS Catalog harmonized files. Signed *Z* scores were computed from effect sizes and standard errors, and the files were processed with munge_sumstats on HapMap3 SNPs [49] excluding the MHC. For BMD the sample size column of the harmonized file was not usable, and the study sample size (426,824) was used for all SNPs. The other nine traits were used as processed LDSC-format files distributed by the Price laboratory: educational attainment and neuroticism [50,51], schizophrenia [52], bipolar disorder [53], BMI [54], IBD [55], rheumatoid arthritis [56], height [35] and LDL cholesterol [36].

### AlphaGenome Atlas predictions

Target SNPs were all biallelic SNPs with MAF ≥ 5% in the 1000 Genomes Phase 3 European reference panel (GRCh38) whose reference and alternative alleles could be matched to the 1000 Genomes high-coverage GRCh38 release (5,828,323 SNPs). We retrieved AlphaGenome Atlas variant scores [18,21] through the AlphaGenome API for the DNase, CAGE, histone ChIP–seq (H3K27ac, H3K4me1, H3K4me3) and RNA-seq scorers, and the overall variant-impact score (AVI). For each track we stored the absolute value of the recommended scorer: the difference between the alternative and reference alleles in the log2 of the predicted signal summed over a 501-bp window centred on the variant (DNase, CAGE) or a 2,001-bp window (histone marks). For RNA-seq, the recommended scorer is a gene-level log fold change, and we took the maximum absolute value over genes. SNPs with MAF < 5% or unresolved alleles were assigned 0 in all annotations.

### Lineage assignment

Tracks were assigned to 12 lineages by ordered regular-expression rules on biosample names and types, using metadata only; the rules were fixed before any heritability analysis. Cell-line tracks were excluded. Biosamples that could not be placed in one lineage were left unassigned; these included mixed blood and bone marrow, germ layers, fetal limbs, peripheral nerve and cerebrospinal fluid. Supplementary Table S2 lists track and biosample counts.

### Annotations

For lineage *g* and layer *m*, the SNP score was the maximum absolute allelic effect across the tracks of *g* and *m*. Scores were converted to percentiles across target SNPs, and the top 5% defined the layer annotation. The lineage annotation (Fig. 2) used the mean of the percentiles for DNase, H3K27ac, H3K4me1 and CAGE, again thresholded at the top 5%. An all-lineage annotation built in the same way from all assigned tracks served as the control in the lineage-specific model. LD scores were computed with a Python 3 port of LDSC [2,57] from the 1000 Genomes European reference (1-cM window) for HapMap3 regression SNPs. On chromosome 22, LD scores computed with this implementation reproduced the published baselineLD v2.2 LD scores exactly.

### Stratified LD score regression

All models contained the 97 annotations of baselineLD v2.2 [3] and used --overlap-annot, allele frequencies for MAF ≥ 5%, HapMap3 regression weights excluding the MHC and a 200-block jackknife. τ* was computed as τ_c_ × sd_c_ × *M* / *h*^2^_g_, where τ_c_ is the per-SNP coefficient of annotation *c*, sd_c_ its standard deviation across common SNPs, *M* the number of common SNPs and *h*^2^_g_ the observed-scale SNP heritability. Unless stated otherwise, *P* values for τ* are one-sided for τ* > 0. Differences between two τ* values in the same model were tested with jackknife standard errors computed from the block delete values.

### Layer map

Each trait was analysed in one lineage, chosen after the lineage map from the lineages enriched for that trait and its expected biology (Supplementary Table S1). For two traits this was not the top-ranked lineage. Bipolar disorder was analysed in brain tissue, although the myeloid lineage ranked first in its small GWAS. Height was analysed in the stromal lineage, which contains chondrocytes, osteoblasts and mesenchymal stem cells, although adipose and skeletal muscle ranked higher. The same lineages were used in the measured-annotation and eQTL comparisons. In the chosen lineage, CAGE, DNase and H3K27ac annotations were fitted jointly. To reduce track-number effects, up to three units per layer were drawn without replacement in each of 20 resamples (seed 20261009). A unit was a track for DNase and H3K27ac and a CAGE library (hCAGE or LQhCAGE), with its two strand-specific tracks, for CAGE; all units were used when three or fewer were available (for example, the two liver CAGE libraries). Estimates and block delete values were averaged over resamples. Because the regression SNPs and jackknife blocks are identical across resamples of a trait, standard errors were computed from the averaged delete values. For each trait, the contrast vector *d* = (CAGE - DNase, CAGE - H3K27ac) was tested for heterogeneity across the 11 traits with a multivariate Cochran’s *Q* for a common mean (20 degrees of freedom; see Tests between traits). Three pairwise contrasts were specified in the protocol: AD versus height and four brain traits (educational attainment, schizophrenia, bipolar disorder and neuroticism) versus height for CAGE - DNase, and IBD and rheumatoid arthritis versus AD and monocyte count for CAGE - H3K27ac (two-sided, α = 0.05/3). Group means were inverse-variance weighted. A trait was assigned a layer type when that layer had the largest resample-averaged τ*, ranked first in at least 15 of 20 resamples and exceeded the second layer with one-sided *P* < 0.05; otherwise it was unresolved.

### Prospective validation

The three validation traits, their analogues, the predicted lineages and the rule that the predicted layer equals the analogue’s layer type were fixed in a dated protocol before the layer map was computed. The resulting predictions were recorded before the validation traits were analysed. For each validation trait we fitted the 12 single-lineage models and the joint three-layer model with the same 20 resamples. The primary test was the one-sided difference in CAGE - DNase between the traits predicted to be CAGE type and those predicted to be DNase type. Secondary tests were enrichment of the predicted lineage (one-sided *P* < 0.05) and confirmation of the predicted layer, which required that layer to exceed both other layers (one-sided *P* < 0.05 for each).

### Threshold sensitivity

Layer annotations were rebuilt with all tracks of each lineage at the top 1%, 0.5% and 0.1% of SNPs, using the same maximum-effect scores and percentiles as the 5% annotations; rebuilding the 5% annotations with the same code reproduced them exactly. The three-layer joint model was fitted once per trait and threshold (42 models), with single-run jackknife standard errors, and the 5% reference was the all-track model of the main analysis. This analysis was specified after the main results had been seen (Supplementary Table S6); its primary criterion at the 0.1% threshold (heterogeneity *P* < 0.05 and a matching top layer in at least 9 of 11 traits, including DNase for height) was fixed in a dated protocol before the analysis.

### Cell-type-matched layer map

Within each analysed lineage, we grouped the biosamples of the three layers that shared an ontology term or that denoted the same cell type or anatomical region under different terms (Supplementary Table S8). From each group we took one unit per layer (one CAGE library with both strands, or one DNase or H3K27ac track), choosing by a fixed order of terms. Each layer therefore had the same number of units in a lineage: one in myeloid (CD14^+^ monocytes), four in brain tissue, three in stromal, two in liver and five in lymphoid. Annotations were built as in the main analysis from the maximum effect over these units and the top 5% of SNPs, and the joint three-layer model was fitted once per trait in its own lineage. The matching table and the primary criterion (an unchanged top layer in at least 7 of the 8 traits with a confirmed type, including DNase for height and CAGE for AD) were fixed in a dated protocol before the analysis.

### Comparison within shared lineages

For each trait we fitted the all-track joint model in every lineage in which the trait was enriched in the lineage map (one-sided *P* < 0.05). In lineages in which both a DNase-type trait (height, BMD) and a CAGE-type trait (AD, monocyte count, educational attainment, schizophrenia, bipolar disorder, BMI, LDL or total cholesterol) were enriched, we computed the difference between the inverse-variance-weighted CAGE - DNase contrasts of the two groups. Differences were pooled across lineages by inverse-variance weighting. Because the same traits contributed to several lineages, the protocol used a conservative standard error that assumed perfect correlation between lineages; we also report the standard error from the joint jackknife, which estimates this correlation (Tests between traits). The criterion (pooled one-sided *P* < 0.05 with the conservative standard error and a positive difference in at least four of six lineages) was fixed in a dated protocol before the analysis. Both control analyses were specified after the main results had been seen (Supplementary Table S6).

### Proximal and distal annotations

TSSs were the start positions of all GENCODE v46 transcripts of protein-coding and lncRNA genes on chromosomes 1–22 (190,108 unique positions) [31]. SNPs within 1,000 bp of a TSS were proximal (5.4% of target SNPs), and the others distal. Each all-track 5% layer annotation of the main map was split into its proximal and distal parts without re-ranking, and the six annotations of a lineage (three layers, each proximal and distal) were fitted jointly with baselineLD v2.2, once per trait in its own lineage, with the joint jackknife across traits. The primary criterion (at distal SNPs, CAGE ranked first for at least 5 of the 7 CAGE-type traits and DNase for height, and heterogeneity *P* < 0.05) was fixed in a dated protocol before the analysis, which was specified after the main results had been seen (Supplementary Table S6). Because τ* depends on annotation size, proximal and distal τ* were not compared with each other.

### Tests between traits

Tests that combined or compared traits used a joint block jackknife. LDSC divides the regression SNPs into 200 blocks of equal size, so block boundaries differ between traits with different SNP sets. We therefore fixed 200 block boundaries by genomic position, dividing the 1,186,879 HapMap3 regression SNPs of the reference weights into blocks of equal size, and refitted every model with these blocks, so that the same genomic segments were deleted for all traits. Point estimates were unchanged. For heterogeneity, the contrasts *d* of all traits were stacked into one vector with its joint jackknife covariance, and *Q* was the generalized least-squares statistic for a common mean (2(*K* - 1) degrees of freedom for *K* traits). We also report an *F* approximation that allows for estimating this covariance from 200 blocks, *F* = (ν - *p* + 1)*Q*/(ν*p*) with *p* = 2(*K* - 1) and ν = 199. Contrasts between groups of traits kept the inverse-variance weights of the main analysis, and their standard errors were computed from the jackknife delete values of the weighted combination, which include the covariance within and between groups. This analysis was specified in a dated protocol after the main results had been seen (Supplementary Table S6); its values replace those computed with traits treated as independent, which are listed in Supplementary Table S10.

### Measured annotations

For each lineage, measured annotations were built for the same ontology terms as the Atlas tracks. For CAGE, we used reprocessed FANTOM5 robust CAGE peaks (phase 1 and 2, GRCh38) and transcribed enhancers [7,8]. FANTOM5 libraries were assigned to a term through the FANTOM5 sample ontology and restricted to libraries of the same sample category (tissue or primary cell) as the Atlas biosample. A peak or enhancer was active for a term when it was expressed (TPM ≥ 1) or used in the majority of that term’s libraries. For DNase and H3K27ac we used ENCODE GRCh38 default peak files [38] (867 files from 683 experiments), and a position was active for a term when it was covered in the majority of the term’s experiments. Active positions were extended by ±250 bp (CAGE, DNase) or ±1,000 bp (H3K27ac) to match the scorer windows, and they were combined over the terms of a lineage. Measured annotations covered 0.5–1.2% (CAGE), 5–23% (DNase) and 12–17% (H3K27ac) of common SNPs. We fitted a same-layer model (baselineLD + measured + predicted annotation of one layer) and a full conditional model (baselineLD + the three measured layers + the predicted annotation). In the primary analysis, specified in the protocol before the comparison was run, the predicted annotation was CAGE for all traits, with false discovery rate control across the 11 traits.

### Comparison with eQTLs

The fine-mapped eQTL annotation of baselineLD (GTEx_eQTL_MaxCPP) [24] was compared with the predicted annotation in the same model through the jackknife difference of their τ* values. Attenuation was the relative change in the eQTL τ* between the baseline-only model and the model with the predicted annotation.

### Robustness analyses for AD

In the initial analysis, the myeloid annotation was based on monocyte and macrophage tracks, scored with the same percentile mean as the lineage annotation and thresholded at 1%, 5% and 10% or used as a continuous percentile score. We also conditioned on the top 5% of the AVI score and repeated the analysis with summary statistics that retained the *APOE* region. In a post hoc analysis, we removed all SNPs within 1 Mb of genome-wide significant variants (*P* < 5 × 10^-8^) and of the lead variants of 75 reported loci, a total of 162 Mb, and we computed leave-one-block-out estimates over the 200 jackknife blocks.

### Analysis protocols and sequence

Each analysis stage was specified in a dated protocol before that stage was run, and all S-LDSC models were run sequentially with fixed software and reference files. Because the stages were sequential, later protocols were written with knowledge of earlier results. For example, the layer-map protocol (9 October 2026) was written after the single-layer map and the joint six-layer models (2–4 October), in which a CAGE cell was the strongest for 10 of 11 traits. Supplementary Table S6 lists, for each stage, the results that had been seen when its protocol was written. Only the predictions for the three validation traits were fixed before any data for those traits were analysed. Post hoc analyses are labelled as such in the text, in Table 1 and in Supplementary Table S6.

## Ethics statement

This study used only publicly available GWAS summary statistics and publicly available functional genomic data, and no individual-level data were accessed. Ethical approval and informed consent were obtained by the original studies, and no separate ethical approval was required for this analysis.

## Data availability

GWAS summary statistics are available from the GWAS Catalog (AD, GCST90027158; monocyte count, GCST90002393; neutrophil count, GCST90002398; BMD, GCST006979; total cholesterol, GCST90500025) and, for the other nine traits, as processed LDSC-format files from the Price laboratory (Supplementary Table S1). AlphaGenome Atlas scores [21] were obtained through the AlphaGenome API. The baselineLD v2.2 annotations, 1000 Genomes reference files and regression weights for GRCh38 are publicly available. FANTOM5 and ENCODE data were obtained from their public portals. Annotation definitions, dated analysis protocols and per-trait results will be made publicly available upon publication, subject to the AlphaGenome terms of use.

## Acknowledgements

We thank the Google DeepMind AlphaGenome team for making the AlphaGenome model and the precomputed AlphaGenome Atlas variant scores available to the research community. We thank the investigators of the European Alzheimer & Dementia Biobank (EADB) consortium and Bellenguez et al., and of the other GWAS consortia whose summary statistics we used, for making their GWAS summary statistics publicly available; the Price laboratory for distributing processed summary statistics and the baselineLD annotations; and the FANTOM5 and ENCODE consortia for their data. The Genotype-Tissue Expression (GTEx) Project, whose data underlie the fine-mapped eQTL annotation of baselineLD, was supported by the Common Fund of the Office of the Director of the National Institutes of Health, and by NCI, NHGRI, NHLBI, NIDA, NIMH and NINDS.

## Funding

This research was supported by a grant of the Korea Health Technology R&D Project through the Korea Health Industry Development Institute (KHIDI), funded by the Ministry of Health & Welfare, Republic of Korea [grant number RS-2025-25455095].

## Author contributions

B.C. designed the study, performed the analyses and wrote the draft of the manuscript. J.J. supervised the study and reviewed the manuscript. J.J. and B.C. revised the manuscript.

## Conflicts of interest

The authors declare no conflicts of interest.

## Notes

### Competing Interest Statement

The authors have declared no competing interest.

### Author Declarations

The study used ONLY openly available human data that were originally located at: - GWAS Catalog (https://www.ebi.ac.uk/gwas/): Alzheimer's disease, GCST90027158; monocyte count, GCST90002393; neutrophil count, GCST90002398; heel bone mineral density, GCST006979; total cholesterol, GCST90500025. - Processed LDSC-format GWAS summary statistics distributed by the Price laboratory (https://alkesgroup.broadinstitute.org/sumstats_formatted/): educational attainment, neuroticism, schizophrenia, bipolar disorder, body mass index, inflammatory bowel disease, rheumatoid arthritis, height and LDL cholesterol. - baselineLD v2.2 annotations, LD reference files and regression weights from the Price laboratory (https://alkesgroup.broadinstitute.org/LDSCORE/). - 1000 Genomes Project reference data (https://www.internationalgenome.org/). - FANTOM5 CAGE data (https://fantom.gsc.riken.jp/5/). - ENCODE DNase-seq and H3K27ac data (https://www.encodeproject.org/).

