## Supplementary Information for "Sequence-based variant-effect predictions reveal trait-specific regulatory-layer profiles of complex-trait heritability"

for

**Contents:** Supplementary Tables S1–S11; Supplementary Figures S1–S3. Reference numbers refer to the reference list of the main text.

### Supplementary Tables

**Supplementary Table S1 | GWAS summary statistics and lineages.** *N* is the median per-SNP sample size after processing (cases plus controls for case–control traits). *h*^2^_g_ is the observed-scale SNP heritability from the S-LDSC models (it varies little between models). Lineage τ* is from the single-lineage model (Fig. 2) for the lineage used in the layer analyses, with its rank among 12 lineages. For the validation traits, the lineage was predicted before analysis.

| **Trait** | **Role** | **Study** | **Source** | ***N*** | ***h*^2^_g_** | **Lineage used** | **Lineage τ* (rank)** |
| --- | --- | --- | --- | --- | --- | --- | --- |
| Alzheimer's disease | Map | Bellenguez 2022 [48] | GCST90027158 (stage I) | 487,511 | 0.026 | Myeloid | +2.98 (1) |
| Educational attainment | Map | Okbay 2016 [50] | Price laboratory | 328,917 | 0.110 | Brain tissue | +0.87 (1) |
| Schizophrenia | Map | PGC 2014 [52] | Price laboratory | 70,100 | 0.458 | Brain tissue | +1.55 (1) |
| Bipolar disorder | Map | PGC 2011 [53] | Price laboratory | 16,731 | 0.390 | Brain tissue | +1.91 (2) |
| Neuroticism | Map | Okbay 2016 [51] | Price laboratory | 170,911 | 0.096 | Brain tissue | +1.42 (1) |
| BMI | Map | Speliotes 2010 [54] | Price laboratory | 123,822 | 0.146 | Brain tissue | +0.53 (1) |
| Monocyte count | Map | Vuckovic 2020 [34] | GCST90002393 | 408,112 | 0.246 | Myeloid | +2.82 (1) |
| IBD | Map | Jostins 2012 [55] | Price laboratory | 34,652 | 0.366 | Lymphoid | +3.43 (1) |
| Rheumatoid arthritis | Map | Okada 2014 [56] | Price laboratory | 38,242 | 0.187 | Lymphoid | +2.48 (1) |
| LDL cholesterol | Map | Teslovich 2010 [36] | Price laboratory | 95,386 | 0.102 | Liver | +3.21 (1) |
| Height | Map | Lango Allen 2010 [35] | Price laboratory | 133,719 | 0.223 | Stromal | +1.54 (3) |
| Bone mineral density (heel) | Validation | Morris 2019 [32] | GCST006979 | 426,824 | 0.285 | Stromal | +1.85 (3) |
| Neutrophil count | Validation | Vuckovic 2020 [34] | GCST90002398 | 408,112 | 0.197 | Myeloid | +2.04 (1) |
| Total cholesterol | Validation | Zoodsma 2025 [33] | GCST90500025 | 434,646 | 0.083 | Liver | +1.97 (1) |

**Supplementary Table S2 | AlphaGenome Atlas tracks per lineage and layer.** Number of tracks and of biosamples (ontology terms with any layer) in each lineage after excluding cell lines (372 tracks) and unassigned biosamples (64 tracks). CAGE tracks are strand specific, two per CAGE library (hCAGE or LQhCAGE); some biosamples have both library types, so that, for example, the 36 myeloid CAGE tracks come from 18 libraries of 13 biosamples.

| **Lineage** | **Biosamples** | **CAGE** | **DNase** | **H3K27ac** | **H3K4me1** | **H3K4me3** | **RNA-seq** | **Examples** |
| --- | --- | --- | --- | --- | --- | --- | --- | --- |
| Brain tissue | 38 | 52 | 12 | 8 | 10 | 11 | 24 | prefrontal and temporal cortex, caudate, hippocampus, cerebellum, spinal cord |
| Neural cells | 17 | 12 | 9 | 6 | 6 | 9 | 9 | cultured neurons, neural progenitors, astrocytes, oligodendrocyte precursors, Schwann cells |
| Myeloid | 17 | 36 | 4 | 3 | 3 | 3 | 2 | CD14^+^ monocytes, macrophages, dendritic cells, granulocytes, mast cells, myeloid progenitors |
| Lymphoid | 32 | 18 | 27 | 13 | 12 | 13 | 30 | T, B and NK cells, thymus, spleen, tonsil |
| Liver | 4 | 4 | 4 | 3 | 3 | 3 | 7 | hepatocytes, liver tissue |
| Gastrointestinal | 46 | 44 | 15 | 19 | 21 | 22 | 31 | oesophagus, stomach, colon, small intestine, pancreas |
| Cardiovascular | 59 | 58 | 29 | 13 | 11 | 14 | 49 | heart, aorta, endothelial cells, vascular smooth muscle |
| Skeletal muscle | 21 | 22 | 12 | 7 | 8 | 9 | 17 | skeletal muscle tissue, myoblasts, myotubes |
| Adipose | 17 | 24 | 3 | 1 | 2 | 2 | 8 | adipose tissue, adipocytes, preadipocytes |
| Stromal/mesenchymal | 67 | 84 | 23 | 7 | 6 | 12 | 27 | fibroblasts, mesenchymal stem cells, chondrocytes, osteoblasts |
| Reproductive/endocrine | 33 | 46 | 10 | 13 | 11 | 11 | 29 | reproductive organs, placenta, thyroid, adrenal gland |
| Epithelial/barrier | 82 | 86 | 40 | 17 | 14 | 22 | 57 | skin, lung and airway, kidney, breast, prostate |
| Total | 433 | 486 | 188 | 110 | 107 | 131 | 290 |  |

**Supplementary Table S3 | Comparison at each trait's own layer.** Own layer: the top layer in the layer map. Atlas alone: τ* of the predicted own-layer annotation conditional on baselineLD. vs eQTL: difference between the predicted and the GTEx eQTL (MaxCPP) τ* in the same model (two-sided *P*). vs measured: difference between the predicted and the measured τ* of the same layer fitted together (two-sided *P*). Conditioned: predicted τ* after conditioning on measured CAGE, DNase and H3K27ac annotations of the same lineage (one-sided *P*; retention, the conditioned τ* as a percentage of the τ* alone). The conditioned model for height was run after the layer map was known (†).

| **Trait** | **Own layer** | **Atlas alone** | **vs eQTL (*P*)** | **vs measured (*P*)** | **Conditioned (*P*; retention)** |
| --- | --- | --- | --- | --- | --- |
| Alzheimer's disease | CAGE | +3.05 | +1.56 (0.065) | +2.91 (0.004) | +2.17 (0.001; 71%) |
| Monocyte count | CAGE | +3.81 | +3.23 (5 × 10^−8^) | +2.28 (0.001) | +2.49 (1 × 10^−7^; 65%) |
| Educational attainment | CAGE | +1.21 | +0.97 (6 × 10^−4^) | +0.63 (0.035) | +0.84 (2 × 10^−4^; 69%) |
| Schizophrenia | CAGE | +1.51 | +0.95 (0.002) | +0.63 (0.093) | +1.07 (4 × 10^−5^; 71%) |
| Bipolar disorder | CAGE | +4.37 | +3.82 (8 × 10^−5^) | +3.55 (0.003) | +4.02 (8 × 10^−7^; 92%) |
| Neuroticism | CAGE | +0.97 | +0.47 (0.22) | +0.14 (0.77) | +0.52 (0.047; 54%) |
| BMI | CAGE | +0.74 | +0.38 (0.32) | +0.28 (0.53) | +0.49 (0.064; 67%) |
| IBD | CAGE | +2.94 | +2.48 (6 × 10^−4^) | +2.42 (0.010) | +2.16 (2 × 10^−5^; 73%) |
| Rheumatoid arthritis | CAGE | +2.95 | +2.80 (8 × 10^−4^) | +2.20 (0.030) | +2.04 (8 × 10^−4^; 69%) |
| LDL cholesterol | CAGE | +4.33 | +4.21 (1 × 10^−5^) | +5.24 (6 × 10^−4^) | +4.23 (3 × 10^−7^; 98%) |
| Height | DNase | +1.78 | +1.64 (5 × 10^−4^) | +0.92 (0.035) | +1.27 (9 × 10^−4^; 72%)† |

**Supplementary Table S4 | Primary test (specified before the comparison): predicted CAGE conditional on measured annotations.** τ* of the predicted CAGE annotation of each trait's lineage, alone and conditional on measured CAGE, DNase and H3K27ac annotations of the same lineage (full model). *q*: Benjamini–Hochberg across 11 traits. Retention: the conditioned τ* as a percentage of the τ* alone. Sensitivity analyses used three CAGE tracks matched in number to the other layers (track-matched) or measured peaks without scorer-window extension (peaks only).

| **Trait** | **Lineage** | **Alone** | **Conditioned (s.e.)** | ***q*** | **Retention** | **Track-matched (*q*)** | **Peaks only (*q*)** |
| --- | --- | --- | --- | --- | --- | --- | --- |
| Alzheimer's disease | Myeloid | +3.05 | +2.17 (0.70) | 0.001 | 71% | +2.69 (< 0.001) | +1.96 (0.003) |
| Monocyte count | Myeloid | +3.81 | +2.49 (0.49) | 2 × 10^−6^ | 65% | +2.37 (< 0.001) | +2.69 (< 0.001) |
| Educational attainment | Brain tissue | +1.21 | +0.84 (0.24) | 4 × 10^−4^ | 69% | +0.80 (< 0.001) | +0.93 (< 0.001) |
| Schizophrenia | Brain tissue | +1.51 | +1.07 (0.27) | 1 × 10^−4^ | 71% | +1.15 (< 0.001) | +1.15 (< 0.001) |
| Bipolar disorder | Brain tissue | +4.37 | +4.02 (0.84) | 3 × 10^−6^ | 92% | +3.70 (< 0.001) | +4.17 (< 0.001) |
| Neuroticism | Brain tissue | +0.97 | +0.52 (0.31) | 0.057 | 54% | +0.22 (0.24) | +0.62 (0.030) |
| BMI | Brain tissue | +0.74 | +0.49 (0.32) | 0.071 | 67% | +0.48 (0.052) | +0.57 (0.044) |
| IBD | Lymphoid | +2.94 | +2.16 (0.53) | 7 × 10^−5^ | 73% | +1.64 (0.002) | +2.11 (< 0.001) |
| Rheumatoid arthritis | Lymphoid | +2.95 | +2.04 (0.64) | 0.001 | 69% | +2.01 (0.003) | +1.86 (0.003) |
| LDL cholesterol | Liver | +4.33 | +4.23 (0.85) | 2 × 10^−6^ | 98% | +4.54 (< 0.001) | +4.20 (< 0.001) |
| Height | Stromal | +0.42 | +0.07 (0.33) | 0.42 | 16% | +0.83 (0.015) | +0.09 (0.40) |

**Supplementary Table S5 | Leave-one-trait-out heterogeneity of layer profiles (post hoc).** Multivariate Cochran's *Q* on the (CAGE − DNase, CAGE − H3K27ac) contrasts of the layer map with one trait removed (18 degrees of freedom; 20 with all 11 traits), computed with the joint jackknife across traits; *P* from the χ^2^ distribution and from the *F* approximation (Methods). Values with traits treated as independent are in Supplementary Table S10.

| **Trait removed** | ***Q*** | ***P*** | ***P* (*F* approximation)** |
| --- | --- | --- | --- |
| None (all 11 traits) | 74.3 | 3.6 × 10^−8^ | 7.6 × 10^−6^ |
| Alzheimer's disease | 72.3 | 1.9 × 10^−8^ | 3.2 × 10^−6^ |
| Monocyte count | 56.3 | 8.0 × 10^−6^ | 1.9 × 10^−4^ |
| Educational attainment | 73.7 | 1.0 × 10^−8^ | 2.2 × 10^−6^ |
| Schizophrenia | 73.9 | 9.9 × 10^−9^ | 2.1 × 10^−6^ |
| Bipolar disorder | 64.5 | 3.8 × 10^−7^ | 2.4 × 10^−5^ |
| Neuroticism | 65.9 | 2.2 × 10^−7^ | 1.7 × 10^−5^ |
| BMI | 71.1 | 3.0 × 10^−8^ | 4.4 × 10^−6^ |
| IBD | 74.3 | 8.5 × 10^−9^ | 1.9 × 10^−6^ |
| Rheumatoid arthritis | 70.0 | 4.5 × 10^−8^ | 5.7 × 10^−6^ |
| LDL cholesterol | 65.7 | 2.4 × 10^−7^ | 1.8 × 10^−5^ |
| Height | 38.5 | 0.0033 | 0.014 |

**Supplementary Table S6 | Analysis sequence and status.** Each stage was specified in a dated protocol before it was run. Because the stages were sequential, each protocol was written with knowledge of the results listed under Results already seen. Prospective: predictions fixed before any data for those traits were analysed. Post hoc: analyses specified after the results they follow up, without a criterion fixed in advance.

| **Stage** | **Protocol date** | **Content** | **Results already seen** | **Status** |
| --- | --- | --- | --- | --- |
| Initial AD analysis | 2026-09-25 | Monocyte/macrophage and other cell-type annotations; thresholds 1%, 5%, 10% and continuous; AVI conditioning; *APOE* region included | Locus-level AD study [22] | Protocol before analysis |
| AD robustness and control traits | 2026-09-28 | Control traits for the myeloid and neuron annotations | Initial AD analysis | Protocol before analysis |
|  | 2026-09-28 | Removal of genome-wide significant loci; leave-one-block-out | Initial AD analysis | Post hoc |
| Lineage map | 2026-09-28 | 12 lineages, 11 traits; AD myeloid, brain traits in brain tissue, AD brain tissue | Initial AD analysis; control traits | Protocol before analysis |
| Single-layer map | 2026-10-02 | 72 lineage–layer annotations; joint six-layer models | Lineage map | Protocol before analysis |
| eQTL comparison | 2026-10-04 | Predicted CAGE versus GTEx MaxCPP in the same model | Lineage and single-layer maps | Post hoc (descriptive) |
| Measured annotations | 2026-10-08 | FANTOM5 and ENCODE annotations; predicted CAGE conditional on three measured layers | Single-layer map; eQTL comparison | Protocol before analysis |
| Layer map with resampling | 2026-10-09 | Three-layer joint models, 20 resamples; heterogeneity, pairwise contrasts, classification | Single-layer map and six-layer models; eQTL and measured-annotation comparisons | Protocol before analysis (exploratory classification) |
| Validation predictions | 2026-10-09 (traits, rule); 2026-10-10 (predictions recorded) | Three new traits; predicted lineage and layer | As above and, when the predictions were recorded, the layer map; no data for the new traits | Prospective |
| Leave-one-trait-out heterogeneity | 2026-10-10 | Supplementary Table S5 | Layer map | Post hoc |
| Own-layer comparison | 2026-10-11 | Supplementary Table S3; one new conditional model for height | Layer map; measured-annotation comparison | Post hoc |
| Threshold sensitivity | 2026-10-11 | Top 1%, 0.5% and 0.1% layer maps; criterion at 0.1% fixed before analysis (Supplementary Table S7) | All of the above | Protocol before analysis |
| Cell-type-matched layer map | 2026-10-13 | One unit per layer from matched cell types or regions (Supplementary Table S8) | All of the above | Protocol before analysis |
| Comparison within shared lineages | 2026-10-13 | All-track joint models in each enriched lineage; skeletal versus lipid and blood traits (Supplementary Table S9) | All of the above | Protocol before analysis |
| Joint jackknife across traits | 2026-10-14 | All layer models refitted with genomic blocks aligned across traits; tests between traits with their covariance (Supplementary Table S10) | All of the above | Protocol before analysis |
| Proximal and distal annotations | 2026-10-15 | Main-map annotations split at 1 kb from the nearest annotated TSS; proximal and distal parts fitted jointly (Supplementary Table S11) | All of the above, including the joint jackknife | Protocol before analysis |

**Supplementary Table S7 | Layer map at stricter annotation thresholds.** Top layer and its τ* in the three-layer joint model (all tracks of the trait's lineage, single run) at each threshold. Asterisk: the top layer exceeds the second with one-sided *P* < 0.05. The 5% column is the all-track version of the main layer map. By the criterion fixed before the analysis (at 0.1%, heterogeneity *P* < 0.05 and a top layer matching the main map in at least 9 of 11 map traits, with DNase for height), the map was not reproduced at 0.1%; at 1% the ranking part was met but heterogeneity was not significant. The summary rows give the tests of the main layer map repeated at each threshold, with the joint jackknife across traits: the pairwise contrasts are two-sided with α = 0.017, and the validation contrast is the one-sided difference in CAGE − DNase between the traits predicted to be CAGE type and DNase type. Brain traits: educational attainment, schizophrenia, bipolar disorder and neuroticism.

| **Trait** | **Top 0.1%** | **Top 0.5%** | **Top 1%** | **Top 5%** |
| --- | --- | --- | --- | --- |
| Educational attainment | DNase +0.58 | CAGE +0.79 | DNase +0.73 | CAGE +1.22* |
| Schizophrenia | CAGE +0.31 | CAGE +0.54 | CAGE +0.39 | CAGE +1.53* |
| Bipolar disorder | H3K27ac +1.89 | CAGE +2.52 | CAGE +2.84 | CAGE +4.79* |
| Neuroticism | DNase +0.62* | DNase +0.47 | DNase +0.59 | CAGE +0.91 |
| BMI | CAGE +0.36 | DNase +0.11 | CAGE +0.57 | CAGE +0.88 |
| Alzheimer's disease | CAGE +3.49 | CAGE +2.26 | CAGE +2.39 | CAGE +2.91* |
| Monocyte count | H3K27ac +1.61 | H3K27ac +2.36 | CAGE +3.11 | CAGE +3.47* |
| IBD | CAGE +2.89 | CAGE +2.72 | CAGE +2.13 | CAGE +2.32 |
| Rheumatoid arthritis | DNase +1.95 | CAGE +2.90 | CAGE +2.36 | CAGE +2.98* |
| LDL cholesterol | DNase +3.67 | CAGE +2.74 | CAGE +3.57 | CAGE +3.91* |
| Height | DNase +0.85 | DNase +0.86 | DNase +1.47 | DNase +1.65* |
| Bone mineral density | DNase +0.96 | DNase +0.98 | DNase +1.47 | DNase +1.51 |
| Neutrophil count | CAGE +1.95* | CAGE +1.74 | CAGE +1.61 | CAGE +1.35 |
| Total cholesterol | DNase +1.57 | CAGE +2.66* | CAGE +2.82 | CAGE +3.70* |
| Summary across the 11 map traits |  |  |  |  |
| Top layer matching the main map | 5 of 11 | 8 of 11 | 9 of 11 | 11 of 11 |
| Traits with top layer exceeding the second | 1 | 0 | 0 | 8 |
| Heterogeneity *Q* (20 d.f.), *P* | 48.1, 4 × 10^−4^ | 40.0, 0.005 | 31.0, 0.055 | 70.1, 2 × 10^−7^ |
| AD vs height, CAGE − DNase, *P* | 0.028 | 0.13 | 0.057 | 0.008 |
| Brain traits vs height, CAGE − DNase, *P* | 0.71 | 0.26 | 0.025 | 8 × 10^−6^ |
| IBD and RA vs AD and monocyte count, CAGE − H3K27ac, *P* | 0.10 | 0.005 | 0.38 | 0.76 |
| Validation contrast, one-sided *P* | 0.018 | 2 × 10^−4^ | 3 × 10^−5^ | 4 × 10^−6^ |

**Supplementary Table S8 | Layer map with cell-type-matched layers.** Three-layer joint models (τ*, standard error) with annotations built from matched cell types or anatomical regions, one unit per layer from each matched group (single run). Matched groups: myeloid, CD14^+^ monocytes (CAGE, CD14^+^CD16^−^ classical monocytes); brain tissue, spinal cord, hippocampus, caudate nucleus and dorsolateral prefrontal cortex (CAGE, middle frontal gyrus); stromal, dermal fibroblasts, lung fibroblasts and mesenchymal stem cells; liver, hepatocytes and liver tissue; lymphoid, natural killer cells, naive CD4^+^ T cells, regulatory T cells, spleen and thymus. Units per layer: myeloid 1, brain tissue 4, stromal 3, liver 2, lymphoid 5. Asterisk: the top layer exceeds the second with one-sided *P* < 0.05. Main map: top layer in the main layer map (validation traits: predicted layer). By the criterion fixed before the analysis (an unchanged top layer in at least 7 of the 8 traits with a confirmed type, including DNase for height and CAGE for AD), the map was retained (8 of 8).

| **Trait** | **Lineage** | **CAGE** | **DNase** | **H3K27ac** | **Top layer** | **Main map** |
| --- | --- | --- | --- | --- | --- | --- |
| Alzheimer's disease | Myeloid | +3.52 (0.84) | +0.83 (0.51) | −0.37 (0.64) | CAGE* | CAGE |
| Monocyte count | Myeloid | +3.78 (0.63) | +1.20 (0.40) | +0.07 (0.41) | CAGE* | CAGE |
| Educational attainment | Brain tissue | +1.28 (0.25) | +0.10 (0.25) | −0.08 (0.23) | CAGE* | CAGE |
| Schizophrenia | Brain tissue | +1.66 (0.28) | −0.05 (0.26) | −0.18 (0.24) | CAGE* | CAGE |
| Bipolar disorder | Brain tissue | +4.93 (0.89) | +2.25 (0.88) | −2.72 (0.88) | CAGE* | CAGE |
| Neuroticism | Brain tissue | +1.06 (0.34) | −0.15 (0.41) | +0.12 (0.33) | CAGE* | CAGE |
| BMI | Brain tissue | +1.09 (0.33) | −0.05 (0.34) | −0.48 (0.34) | CAGE* | CAGE |
| IBD | Lymphoid | +2.35 (0.54) | +0.51 (0.67) | +1.29 (0.73) | CAGE | CAGE |
| Rheumatoid arthritis | Lymphoid | +2.95 (0.80) | +0.32 (0.74) | −0.26 (0.84) | CAGE* | CAGE |
| LDL cholesterol | Liver | +3.93 (0.82) | +0.60 (0.75) | +1.16 (0.71) | CAGE* | CAGE |
| Height | Stromal | +0.16 (0.43) | +2.08 (0.49) | +0.34 (0.40) | DNase* | DNase |
| Bone mineral density | Stromal | +0.18 (0.32) | +2.41 (0.43) | +0.54 (0.40) | DNase* | DNase |
| Neutrophil count | Myeloid | +2.14 (0.41) | +0.83 (0.34) | +0.27 (0.33) | CAGE* | CAGE |
| Total cholesterol | Liver | +3.75 (0.88) | +0.41 (0.45) | −0.10 (0.73) | CAGE* | CAGE |

**Supplementary Table S9 | Comparison of skeletal and lipid or blood traits within shared lineages.** All-track three-layer joint models were fitted for each trait in every lineage in which it was enriched in the lineage map (one-sided *P* < 0.05). Shown are the lineages in which both a DNase-type trait and a CAGE-type trait were enriched; within each lineage all traits share the same annotations. Values are inverse-variance-weighted CAGE − DNase contrasts (τ*, standard error) of the enriched traits of each type. Differences, their standard errors and *P* values use the joint jackknife across traits and lineages; the group values use the standard errors of each trait. With the conservative standard error of the protocol, which assumed perfect correlation between lineages, the pooled difference was +2.29 (0.84), one-sided *P* = 0.003, and by the criterion fixed before the analysis (pooled one-sided *P* < 0.05 with this standard error and a positive difference in at least four of six lineages), the difference was retained. Top layers depended on the lineage: H3K27ac ranked first for height in the adipose, cardiovascular, epithelial/barrier and skeletal muscle lineages, and for LDL or total cholesterol in the gastrointestinal, adipose, cardiovascular, lymphoid and reproductive/endocrine lineages.

| **Lineage** | **CAGE-type traits** | **CAGE − DNase** | **DNase-type traits** | **CAGE − DNase** | **Difference** | **One-sided *P*** |
| --- | --- | --- | --- | --- | --- | --- |
| Reproductive/endocrine | LDL, total cholesterol | +2.35 (0.71) | Height, BMD | −1.58 (0.35) | +3.94 (0.87) | 3 × 10^−6^ |
| Liver | LDL, total cholesterol | +3.29 (0.72) | BMD | +0.44 (0.51) | +2.85 (0.90) | 8 × 10^−4^ |
| Gastrointestinal | LDL, total cholesterol | +2.35 (0.85) | BMD | −0.31 (0.56) | +2.66 (1.12) | 0.009 |
| Epithelial/barrier | LDL | +2.51 (1.10) | Height, BMD | −0.08 (0.35) | +2.58 (1.09) | 0.009 |
| Cardiovascular | Total cholesterol | +1.62 (0.91) | Height, BMD | −0.57 (0.37) | +2.19 (0.98) | 0.013 |
| Adipose | Monocyte count, LDL, total cholesterol | +1.10 (0.50) | Height, BMD | +0.32 (0.38) | +0.78 (0.68) | 0.13 |
| Pooled |  |  |  |  | +2.29 (0.63) | 1.5 × 10^−4^ |

**Supplementary Table S10 | Tests between traits with traits treated as independent and with the joint jackknife.** Original: jackknife blocks of equal SNP number within each trait, traits treated as independent (the values of earlier versions). Aligned: genomic blocks aligned across traits, traits treated as independent. Joint: aligned blocks with the covariance between traits (the values reported in the text). Contrasts are two-sided except the validation contrast and the shared-lineage difference (one-sided). Brain traits: educational attainment, schizophrenia, bipolar disorder and neuroticism. Error correlations of the CAGE − DNase contrast between the 14 traits had a median of +0.03 (range −0.15 to +0.39; largest for monocyte and neutrophil counts, +0.39, and LDL and total cholesterol, +0.30).

| **Analysis** | **Test** | **Original** | **Aligned** | **Joint** |
| --- | --- | --- | --- | --- |
| Layer map (Fig. 3) | Heterogeneity *Q* (20 d.f.), *P* | 66.2, *P* = 7.4 × 10^−7^ | 62.3, *P* = 3.2 × 10^−6^ | 74.3, *P* = 3.6 × 10^−8^ (*F*: 7.6 × 10^−6^) |
|  | AD vs height, CAGE − DNase | +4.51 ± 1.47, *P* = 0.0022 | ± 1.50, *P* = 0.0026 | ± 1.51, *P* = 0.0028 |
|  | Brain traits vs height, CAGE − DNase | +2.75 ± 0.55, *P* = 6.6 × 10^−7^ | ± 0.58, *P* = 1.7 × 10^−6^ | ± 0.58, *P* = 2.4 × 10^−6^ |
|  | IBD and RA vs AD and monocyte count, CAGE − H3K27ac | −1.88 ± 0.83, *P* = 0.023 | ± 0.85, *P* = 0.026 | ± 0.90, *P* = 0.037 |
| Validation (Fig. 4) | CAGE-type vs DNase-type predictions, CAGE − DNase | +3.42 ± 0.70, *P* = 5.1 × 10^−7^ | ± 0.70, *P* = 4.6 × 10^−7^ | ± 0.67, *P* = 1.6 × 10^−7^ |
| Thresholds (Supplementary Table S7) | Heterogeneity, top 0.1% | 41.4, *P* = 0.0033 | 40.9, *P* = 0.0038 | 48.1, *P* = 4.2 × 10^−4^ (*F*: 0.0039) |
|  | Heterogeneity, top 0.5% | 33.2, *P* = 0.032 | 33.6, *P* = 0.029 | 40.0, *P* = 0.005 (*F*: 0.022) |
|  | Heterogeneity, top 1% | 30.4, *P* = 0.064 | 29.7, *P* = 0.075 | 31.0, *P* = 0.055 (*F*: 0.13) |
|  | Heterogeneity, top 5% (all tracks) | 61.9, *P* = 3.7 × 10^−6^ | 58.6, *P* = 1.2 × 10^−5^ | 70.1, *P* = 1.7 × 10^−7^ (*F*: 2.1 × 10^−5^) |
| Cell-type-matched (Supplementary Table S8) | Heterogeneity | 56.8, *P* = 2.2 × 10^−5^ | 53.4, *P* = 7.0 × 10^−5^ | 56.3, *P* = 2.6 × 10^−5^ (*F*: 5.9 × 10^−4^) |
|  | AD vs height | +4.62 ± 1.29, *P* = 3.3 × 10^−4^ | ± 1.32, *P* = 4.8 × 10^−4^ | ± 1.34, *P* = 5.7 × 10^−4^ |
|  | Brain traits vs height | +3.36 ± 0.77, *P* = 1.4 × 10^−5^ | ± 0.81, *P* = 3.2 × 10^−5^ | ± 0.81, *P* = 3.4 × 10^−5^ |
|  | Validation contrast | +4.17 ± 0.74, *P* = 8.6 × 10^−9^ | ± 0.73, *P* = 5.9 × 10^−9^ | ± 0.76, *P* = 1.9 × 10^−8^ |
| Shared lineages (Supplementary Table S9) | Pooled difference (original: conservative standard error of the protocol) | +2.29 ± 0.84, *P* = 0.0033 | – | ± 0.63, *P* = 1.5 × 10^−4^ |

**Supplementary Table S11 | Layer map at distal and proximal SNPs.** Each all-track 5% layer annotation of the main map was split at 1 kb from the nearest TSS of a GENCODE v46 protein-coding or lncRNA transcript, and the proximal and distal parts of the three layers were fitted jointly (τ*, standard error; single run per trait in its own lineage). Distal annotations covered 4.0–4.2% (CAGE), 4.6–4.8% (DNase) and 4.4–4.6% (H3K27ac) of common SNPs and proximal annotations 0.2–1.0%. Asterisk: the top layer exceeds the second with one-sided *P* < 0.05. Across the 11 map traits (joint jackknife), the distal layer profiles differed between traits (*Q* = 63.7, 20 d.f., *P* = 1.9 × 10^−6^; *F* approximation 1.0 × 10^−4^), with CAGE − DNase contrasts of AD versus height of +4.41 (1.49), *P* = 0.003, and of the brain traits versus height of +2.59 (0.65), *P* = 7 × 10^−5^, and a validation contrast of +3.51 (0.70), one-sided *P* = 2 × 10^−7^. The proximal profiles did not differ (*Q* = 18.7, *P* = 0.54). By the criterion fixed before the analysis (at distal SNPs, CAGE first for at least 5 of the 7 CAGE-type traits, DNase for height and heterogeneity *P* < 0.05), the layer map was retained at distal SNPs.

| **Trait** | **Lineage** | **CAGE, distal** | **DNase, distal** | **H3K27ac, distal** | **Top layer, distal** | **Top layer, proximal** |
| --- | --- | --- | --- | --- | --- | --- |
| Alzheimer's disease | Myeloid | +2.86 (0.86) | +0.14 (0.76) | +0.23 (0.58) | CAGE* | CAGE |
| Monocyte count | Myeloid | +2.63 (0.53) | −0.05 (0.46) | +1.40 (0.43) | CAGE* | CAGE* |
| Educational attainment | Brain tissue | +0.94 (0.23) | +0.29 (0.25) | −0.17 (0.23) | CAGE* | CAGE |
| Schizophrenia | Brain tissue | +1.44 (0.26) | +0.19 (0.27) | −0.23 (0.26) | CAGE* | H3K27ac |
| Bipolar disorder | Brain tissue | +4.45 (0.92) | +2.41 (0.87) | −2.64 (0.92) | CAGE* | CAGE* |
| Neuroticism | Brain tissue | +0.83 (0.35) | +0.19 (0.37) | +0.07 (0.34) | CAGE | CAGE |
| BMI | Brain tissue | +0.96 (0.37) | +0.21 (0.39) | −0.55 (0.38) | CAGE | CAGE |
| IBD | Lymphoid | +1.78 (0.56) | +1.21 (0.71) | +0.70 (0.65) | CAGE | CAGE |
| Rheumatoid arthritis | Lymphoid | +2.66 (0.80) | +0.99 (0.89) | −0.42 (0.79) | CAGE | CAGE |
| LDL cholesterol | Liver | +4.24 (0.92) | +0.67 (0.84) | +0.62 (0.65) | CAGE* | DNase |
| Height | Stromal | −0.18 (0.37) | +1.50 (0.44) | +0.30 (0.42) | DNase | DNase |
| Bone mineral density | Stromal | +0.11 (0.30) | +1.46 (0.35) | +1.04 (0.33) | DNase | DNase |
| Neutrophil count | Myeloid | +1.18 (0.45) | +0.16 (0.42) | +1.08 (0.35) | CAGE | DNase |
| Total cholesterol | Liver | +3.61 (0.78) | −0.06 (0.51) | +0.11 (0.66) | CAGE* | DNase |

### Supplementary Figures

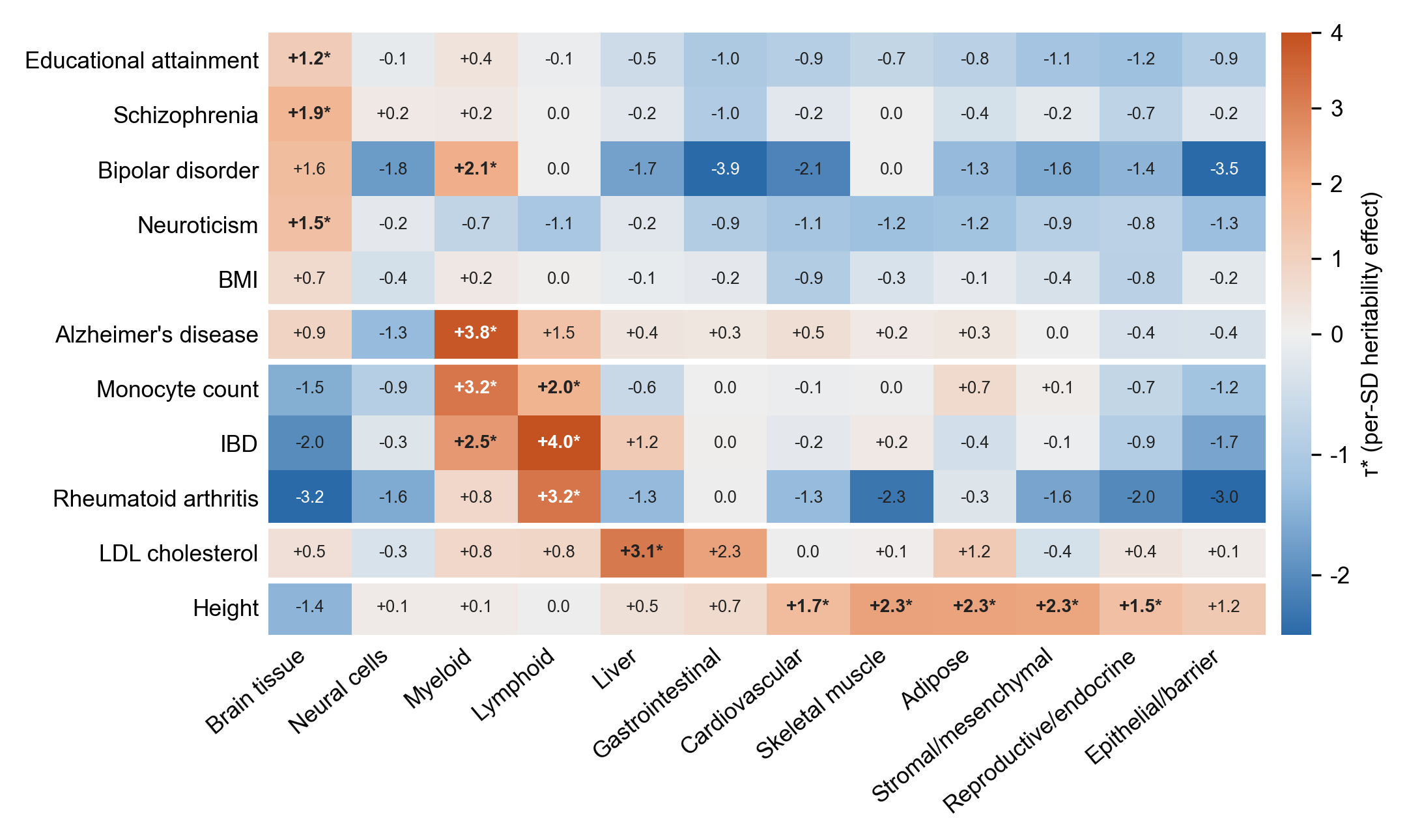

**Supplementary Figure S1 | Lineage-specific heritability enrichment.** τ* of each lineage annotation in models that also contain an all-lineage annotation of SNPs with large predicted effects in any lineage (baselineLD + all-lineage + one lineage annotation). Bold values with an asterisk: false discovery rate < 0.05 across the 132 cells.

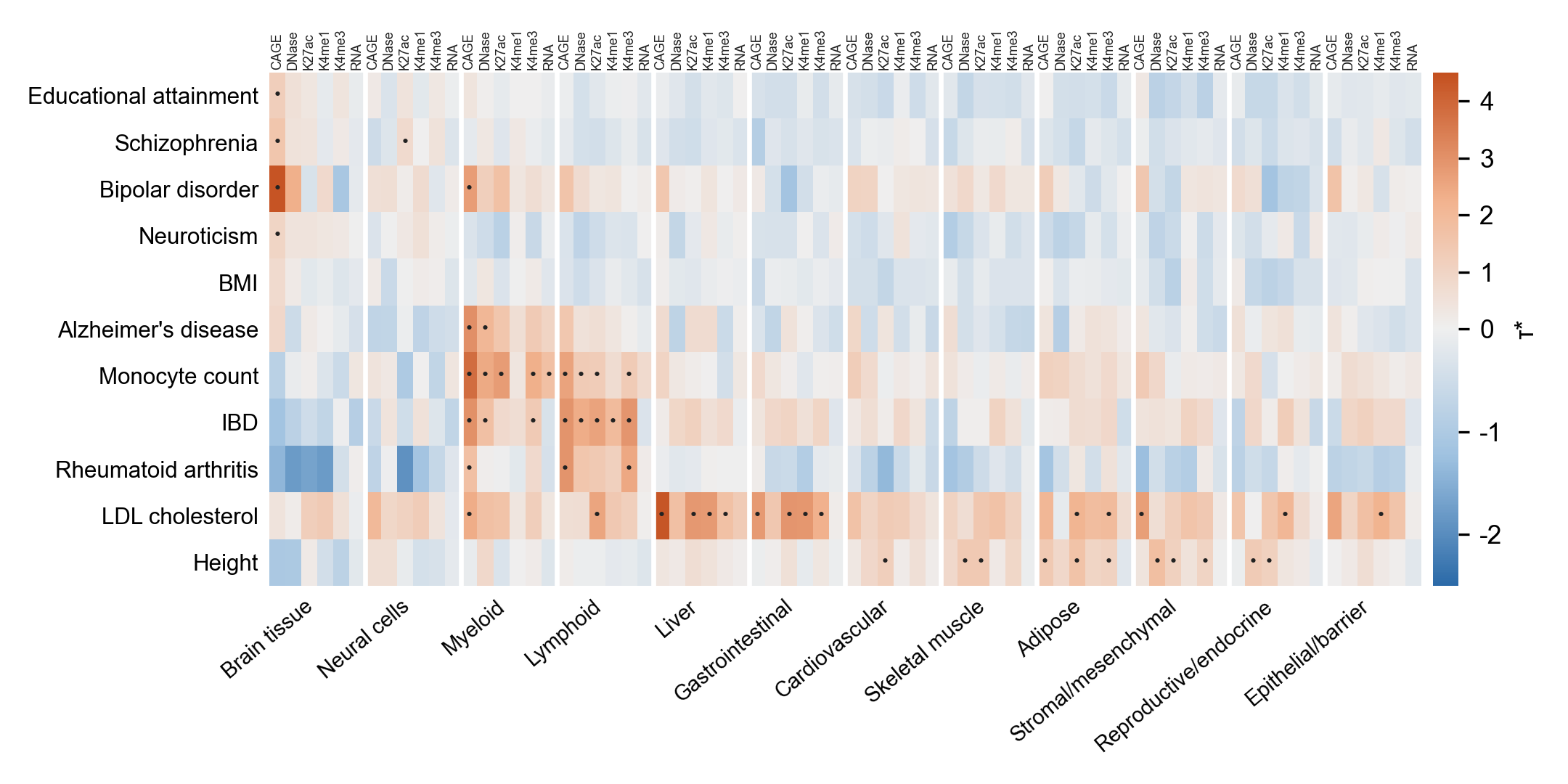

**Supplementary Figure S2 | Single-layer annotations by lineage.** τ* of the 72 lineage–layer annotations (six layers within each of 12 lineages), each fitted alone with baselineLD v2.2. Layers are labelled above the columns (K27ac, K4me1 and K4me3: H3K27ac, H3K4me1 and H3K4me3). Dots: false discovery rate < 0.05 across the 792 cells (54 cells).

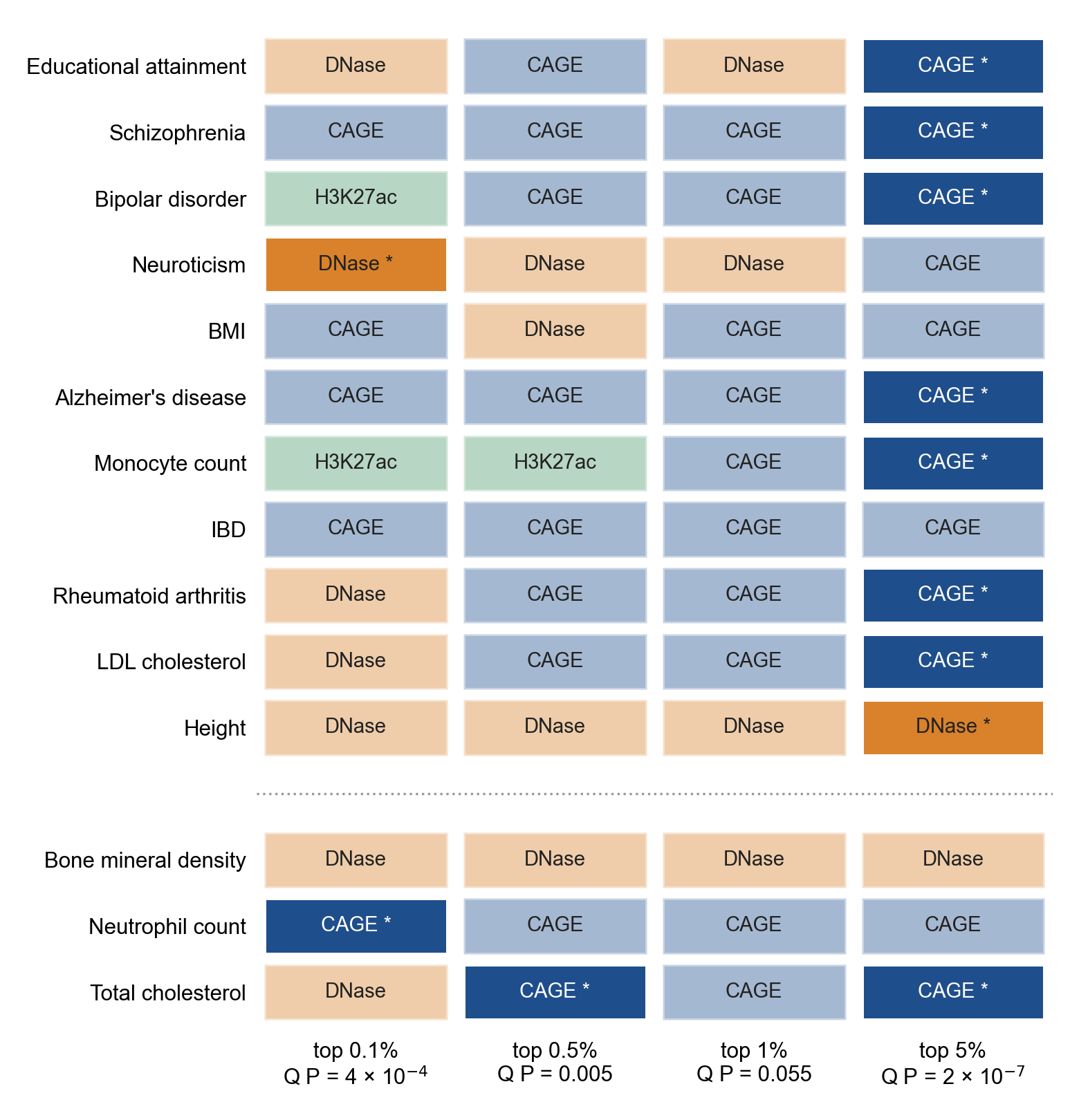

**Supplementary Figure S3 | Layer map at stricter annotation thresholds.** Top layer of each trait in the three-layer joint model at the top 0.1%, 0.5%, 1% and 5% of SNPs (all tracks). Dark cells with an asterisk: the top layer exceeds the second with one-sided *P* < 0.05. The heterogeneity *P* across the 11 map traits (joint jackknife) is shown under each threshold. Validation traits are below the dotted line.
